# Disease-anchored organ aging clocks enable multi-organ risk profiling from routine labs

**DOI:** 10.64898/2026.11.22.26368688

**Authors:** Chang Shi, Yao Sun, Xiaohao Ruan, Ruofan Shi, Chi Yao, Hantang Zhou, Xiangchuan Xu, Xue Li, Rong Na

## Abstract

Organ clocks estimate the biological age of organs from blood, but current clocks assign tissue-enriched proteins to organs and train on chronological age, which requires proteomics and does not yield organ-specific disease risk. In the UK Biobank, we trained organ clocks on first onset of disease of each organ using routine laboratory tests and froze the coefficients. Under an age label, organ-clock readouts arose equally in size-matched random protein panels; with a disease label, clocks selected their own organ’s proteins and predicted their own organ’s diseases. Defining extreme aging by multiples of disease risk concentrated flags on own-organ disease. Frozen clocks kept organ correspondence in the US NHANES cohort, predicted first onset of own-organ disease in the Japanese JMDC cohort and added more information to clinical risk scores than current organ clocks. Trained once where disease follow-up exists, organ clocks can be used wherever routine laboratory tests are measured.

## Introduction

Routine health checks measure a panel of blood tests in large numbers of adults every year. The risk scores used to read these tests each look at one organ, such as the Fibrosis-4 (FIB-4) index for liver fibrosis^1^ and glycated haemoglobin (HbA1c) for diabetes risk. Each score has an inconclusive range, 1.30 to 2.67 for FIB-4^2^ and 42 to 47 mmol/mol for HbA1c^3^. People who fall in it need further tests or a repeat test later. Whether one set of routine tests can give disease risk organ by organ, and pick out people at high risk within these ranges, is a question organ clocks could answer.

Organ-specific aging clocks estimate the biological age of organs from circulating molecular measurements. Existing plasma organ clocks assign features to specific organs using tissue enrichment datasets, train regression models against chronological age, and interpret the residual (the difference between predicted and chronological age) as organ-specific age acceleration or deceleration. Each individual thus receives an organ-resolved panel of biological age gaps. Oh et al.^4^ developed aging clocks for 11 organs using the SomaScan plasma proteome. Goeminne et al.^5^ and Wang et al.^6^ established similar clocks on the Olink proteome of the UK Biobank. Organ-level clocks have since been extended to cell types in the mouse brain^7^, to histology of 40 tissues^8^, to multi-organ systems in the nuclear magnetic resonance (NMR) metabolome^9^ and to sex-stratified multi-omic organ clocks^10^. These age gaps are used as risk markers for disease and death of the organs, to stratify risk by organs^11^ and to classify people by their oldest organs.

The validity of these applications rests on the fundamental assumption that assigning biomolecules to organs by tissue expression confers true organ specificity at the individual level. Prior studies support this assumption with three main lines of evidence: (1) organ age gaps correlate weakly with one another, interpreted as organ autonomy; (2) organ scores associate more strongly with cognate organ diseases than do whole-body clocks or non-target organ clocks; and (3) applying a fixed threshold identifies a distinct subset of individuals with at least one extremely aged organ.

Critically, all three observations can arise as statistical artifacts in the absence of biological organ specificity. First, every organ clock is trained on chronological age, and the age gap represents an ordinary regression residual. Residuals derived from non-overlapping feature sets exhibit low covariance by construction, regardless of whether the features reflect distinct organ biology. Second, comparisons against whole-body clocks are affected by model capacity, as whole-body clocks aggregate one to two orders of magnitude more features. A cognate organ–disease association demonstrates true specificity only if its effect size significantly exceeds that of randomly selected feature panels of the same size. Third, defining extreme aging as an age gap exceeding a fixed parametric threshold (for example 2 standard deviations, SD) fixes the proportion of flagged individuals purely by statistical definition, irrespective of underlying disease risk. Applying an identical cut-off across all organs overlooks the empirical reality that baseline disease incidence varies by two orders of magnitude across organ systems^12^. We therefore test each of the three against random panels.

To overcome these limitations, organ-specific information should be encoded directly into the supervised training objective rather than relegated to residual variation. Paralleling second-generation epigenetic clocks that improved phenotypic capture by training on mortality or biomarker composites^13,14^, we hypothesized that organ identity should be defined by organ-specific clinical endpoints. Specifically, we replace chronological age with the first onset of organ-specific disease (organ-specific morbidity, OS-Morb) or death attributable to that organ (organ-specific mortality, OS-Mort). Using a factorial design, we systematically crossed training labels with feature provision strategies (organ-enriched versus unassigned feature pools).

Across 52,614 UK Biobank participants^15^, we demonstrate that chronological age clocks built from organ-enriched proteomic panels provide negligible organ specificity beyond size-matched random panels. True organ specificity emerges only when models are optimized directly on organ disease outcomes. Furthermore, when extreme aging is redefined according to 5-year absolute disease risk rather than arbitrary standard deviations, flagged individuals concentrate selectively in target-organ pathologies, and each flag corresponds to an explicit risk multiple. Finally, we demonstrate that OS-Morb clocks train effectively on routine laboratory tests and transfer reliably to the external US National Health and Nutrition Examination Survey (NHANES) cohort^16^ and to the Japanese Medical Data Center (JMDC) cohort^17^. Organ clocks can therefore be computed from the laboratory tests of a routine health check and give disease risk arranged by organ.

## Results

### Under an age label, organ-specific clock readouts appear equally on random protein panels

An organ clock is built from two choices: feature allocation and the training objective (Fig. 1a). Features can either be assigned a priori via Human Protein Atlas (HPA) tissue enrichment^18^ or pooled without assignment across all organs (913 features). Training labels can be chronological age (yielding conventional age acceleration, AA), incident organ-specific morbidity (OS-Morb), or organ-specific mortality (OS-Mort)^19^ (Supplementary Table 1). For a given feature set, pipelines differ only in the training label while keeping model architecture, cross-validation, scaling and other steps identical (Fig. 1b). We audited existing approaches in an Olink proteomic cohort (n = 52,614), evaluated factorial combinations across 25 organ–disease pairs in a multi-omics cohort (n = 22,689), and tested translation to routine laboratory tests (n = 390,087) and to external cohorts, the US NHANES cohort (n = 48,916; 7,565 deaths) and the Japanese JMDC health check-up and claims cohort (n = 3,190,470) (Fig. 1).

**Fig. 1.**
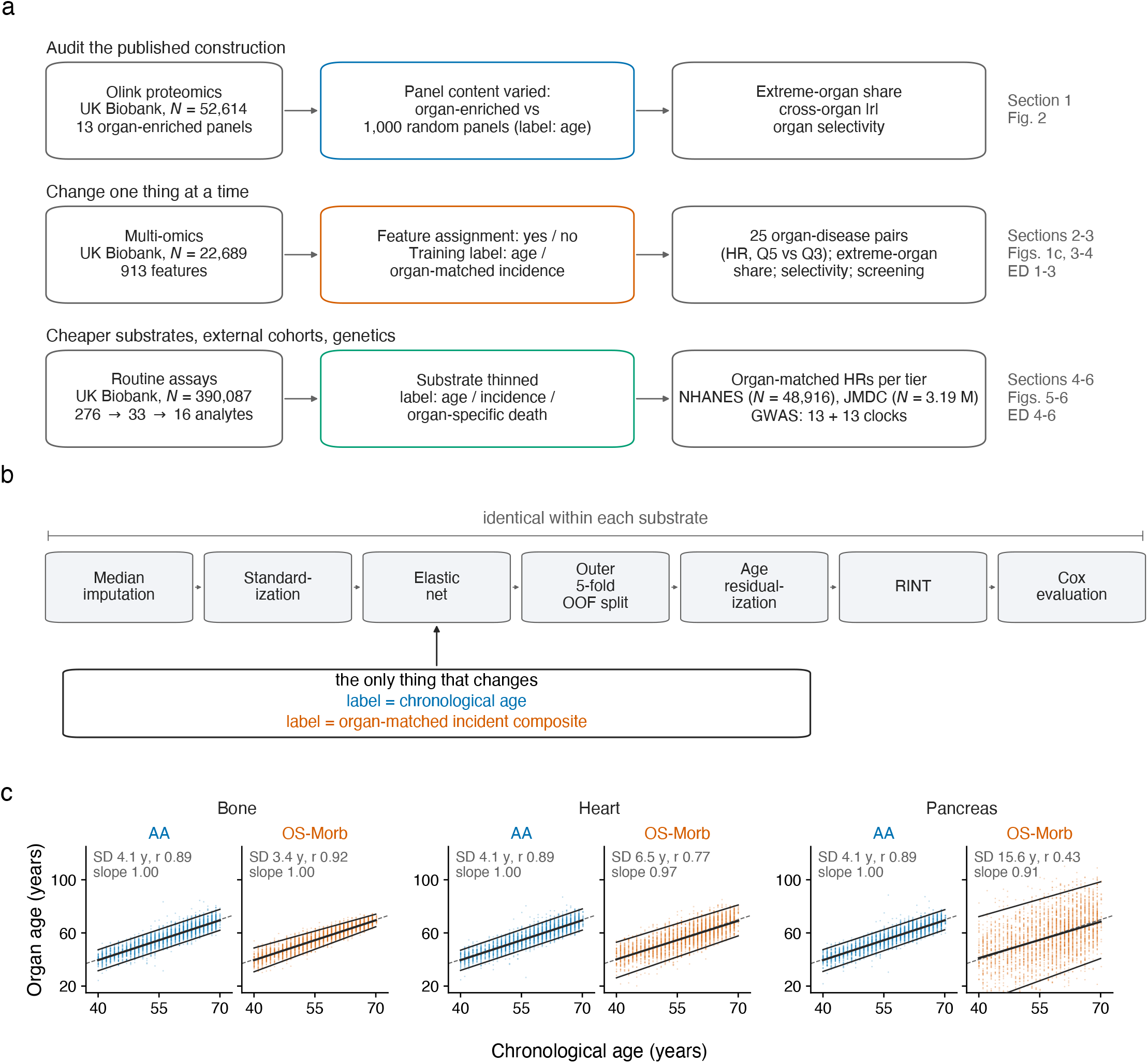
| Study design, and organ age under the two labels. **a**, The three data layers and the element changed in each. **b**, The training pipeline from AA to OS-Morb: the seven steps are identical and only the training label changes. c, Organ age against chronological age for bone, heart and pancreas, from the AA clock (chronological age plus its age gap) and from the OS-Morb score converted to years; multi-omics cohort, n = 22,689, 3,000 random people shown; solid lines, mean and 2.5th and 97.5th percentiles in each year of age, each joined by a straight line; dashed line, y = x; SD, standard deviation of organ age minus chronological age; slope, slope of the means on chronological age.

Tissue-enriched protein panels and size-matched random panels yielded nearly indistinguishable inter-clock correlations under chronological age training. The mean pairwise correlation among the 13 enriched organ-specific proteomic clocks was r = 0.27, falling well within the null distribution generated from 1,000 random panels (median r = 0.28, 2.5th–97.5th percentiles 0.23–0.32; Fig. 2a). Thus, low between-clock correlation is an expected mathematical property of sparse feature partitioning rather than evidence of organ autonomy. Similarly, organ selectivity, defined as the relative risk (RR) for target-organ diseases divided by the RR for non-target organ diseases, was marginal under chronological age training. Enriched panels exceeded size-matched random panels in only 2 of 13 organs (heart and immune system), and the published schemes of Oh et al.^20^, Goeminne et al.^5^ and Wang et al.^6^ in 4 of 10, 3 of 10 and 1 of 9 organs, respectively (Fig. 2b). For example, of the two kidney associations reported by Oh et al.^4^, only one showed discriminative performance above the random-panel distribution (Fig. 2c).

**Fig. 2.**
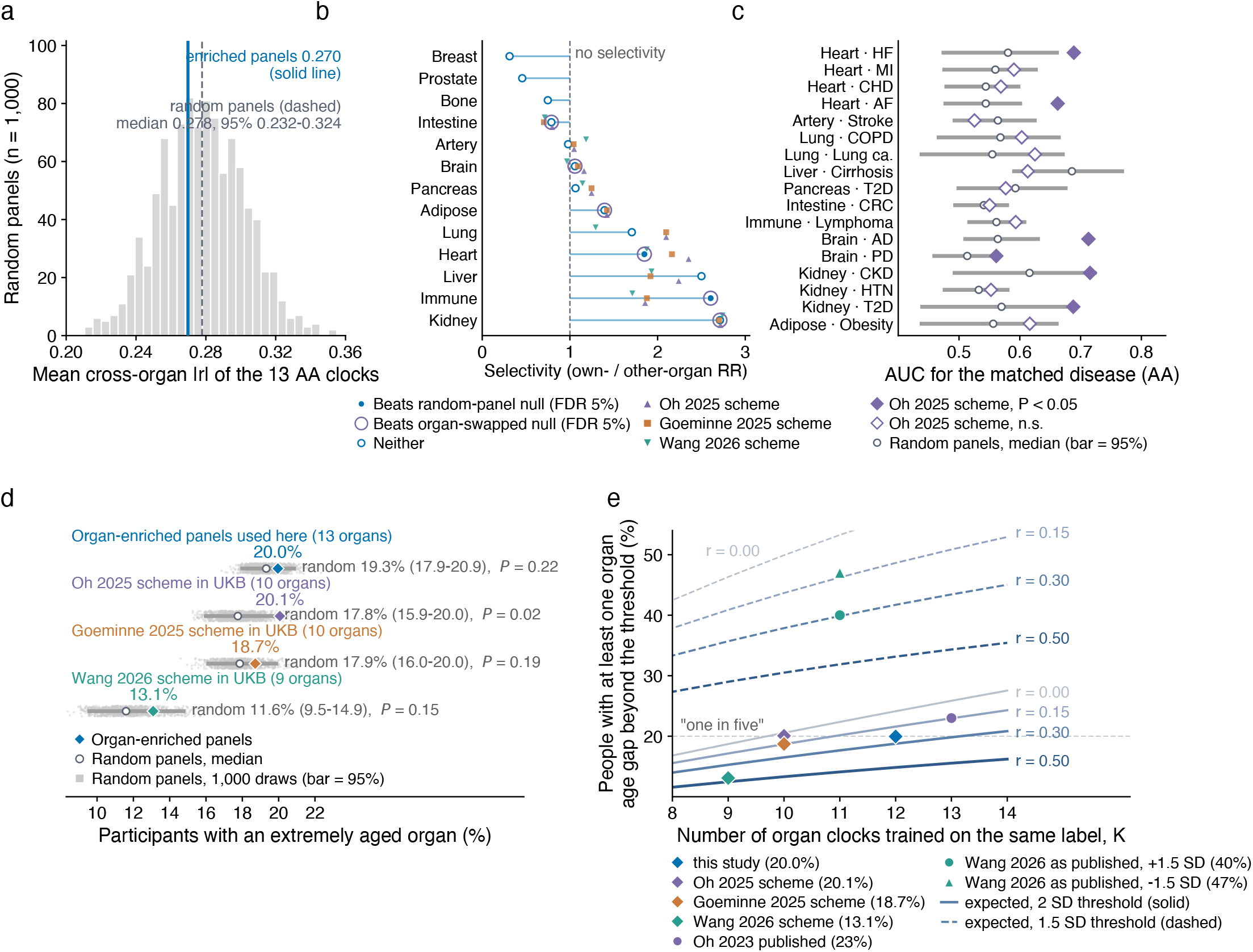
| Under an age label, organ-clock readouts appear equally on random protein panels. Olink proteomic cohort, n = 52,614. **a**, Mean pairwise score correlation among 13 AA organ clocks; histogram, 1,000 random panels of equal size; solid line, enriched panels; dashed line, median of random panels. **b**, Selectivity by organ (own-organ disease RR / other-organ disease RR). Random-panel and organ-swap null models, 1,000 sets each; significance by Benjamini– Hochberg FDR 5%. **c**, AUC on 17 organ–disease pairs under the scheme of Oh et al.^20^, against 1,000 random panels of equal size. Bars, 2.5th to 97.5th percentiles of random panels; P, proportion of random panels not below the observed value. **d**, Proportion of the cohort with an extremely aged organ (any organ score > 2 SD), one row for our panels and one for each published scheme, 1,000 random panels each. **e**, Arithmetic expectation of the extreme-organ proportion as a function of the number of clocks K and the between-clock correlation r (equicorrelated Gaussian model; solid, 2 SD threshold; dashed, 1.5 SD threshold); points, observed or originally reported values of each construction, placed at their own K.

Furthermore, the proportion of individuals flagged with an extremely aged organ (any organ age gap above 2 SD) is set mainly by the number of clocks (K) and their mutual correlation (r), independent of underlying biology. In the proteomic cohort, the observed proportion was 20.0% (95% CI 19.6–20.3%), matching the theoretical multivariate normal expectation of 19.3% and the empirical random-panel null of 19.3% (17.9–20.9% across 1,000 random panels; Fig. 2d,e). Across published schemes, extreme-aging proportions closely mirrored their respective random-panel nulls (Fig. 2d, Supplementary Table 2).

### Organ specificity is driven predominantly by the training label

When the training objective was switched from chronological age to incident organ disease (OS-Morb), organ clocks separated robustly regardless of feature pre-selection. Under chronological age training with organ-assigned proteins, the median pairwise correlation between clocks was 0.63 (95% CI 0.53–0.76). Under OS-Morb, this correlation dropped to 0.34 (95% CI 0.22–0.50) without feature assignment and 0.29 (95% CI 0.20–0.40) with protein assignment (Fig. 3a, Supplementary Table 3). However, this phenotypic separation could not reflect genuine biological selectivity; rather, it was mainly influenced by the selected features in the models. When all 913 multi-omic features were made available to every clock without a priori assignment, OS-Morb models autonomously selected proteins enriched in their cognate organ: the 50 proteins with the largest coefficients in each clock showed a 2.37-fold enrichment (95% CI 1.54–4.37) for target-organ proteins over chance expectation, whereas AA models showed no enrichment (0.94-fold, 95% CI 0.82–2.19; Fig. 3b). Under chronological age labels, every organ clock collapsed onto the same generalized proteomic signature of systemic aging.

**Fig. 3.**
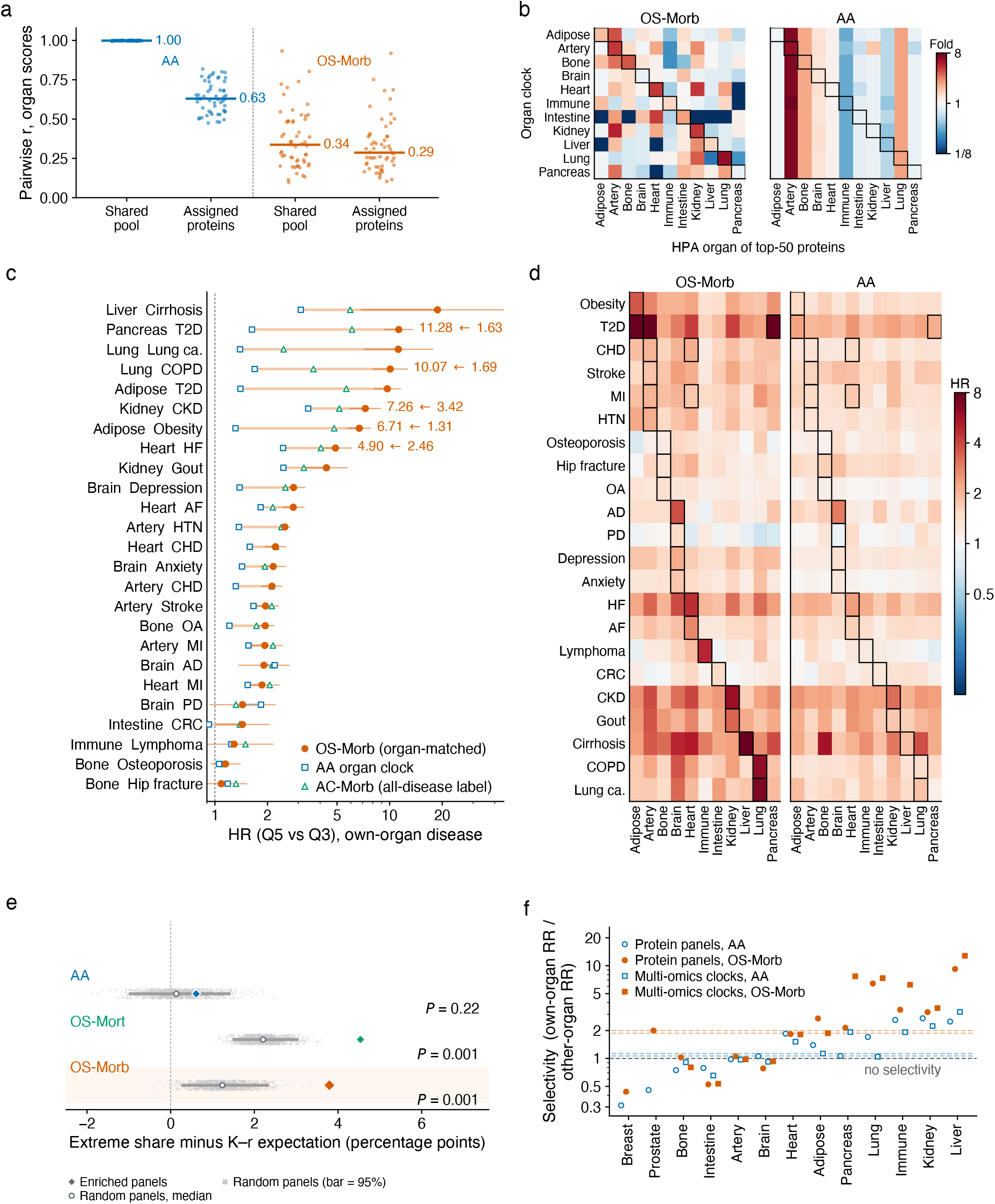
| Changing only the training label brings organ specificity. Multi-omics cohort, n = 22,689. Olink panels in **e** and **f** are from the Olink cohort, n = 52,614. **a**, Pairwise score correlation among 11 organ clocks present in both sexes in the four combinations of feature provision and training label; 55 pairs per combination; horizontal lines, medians. **b**, With features not assigned, the share of each HPA organ among the 50 proteins with the largest absolute coefficients in each organ clock, divided by the chance expectation (log colour scale); boxes, own organ. **c**, Q5 versus Q3 Cox hazard ratios for 25 organ–disease pairs (log axis, adjusted for age and sex); lines, 95% CI. **d**, Q5 versus Q3 Cox hazard ratios of each clock for 22 organ diseases (log colour scale), each disease counted only among people free of it at baseline; boxes, each clock’s own diseases. **e**, Proportion of people with an extremely aged organ minus the expectation from the number of clocks and the between-clock correlation (percentage points), one row per training label, enriched panels against 1,000 random panels. **f**, Selectivity by organ (log axis); dashed lines, medians of each version.

OS-Morb clocks demonstrated superior disease discrimination compared to AA clocks and a pan-disease control clock trained on a composite of all 22 diseases (AC-Morb). Across 25 organ–disease pairs, the hazard ratio comparing the top to the middle quintile (HR Q5 versus Q3) was higher for OS-Morb than for AA trained on the same features in 22 pairs, and higher than for AC-Morb in 18 pairs (Fig. 3c, Supplementary Table 4). In the hazard-ratio matrix over 22 diseases, effect sizes for OS-Morb clustered strongly along the diagonal (geometric mean HR 3.07 for cognate diseases versus 1.69 for non-cognate diseases), whereas AA clocks showed diffuse associations across all endpoints (geometric mean HR 1.59 versus 1.47; Fig. 3d).

Under clinical disease labels, extreme-aging metrics reflected biological risk rather than pure multivariate geometry. Under OS-Morb, even the median random panel exceeded the expectation from the number of clocks and the between-clock correlation (K-r expectation), by 1.2 percentage points (95% CI 0.3–2.4); the proportion of flagged individuals on enriched panels exceeded it by 3.8 percentage points (95% CI 3.5–4.0), more than random panels (P = 0.001). The same pattern held under organ-specific mortality labels (OS-Mort). Under chronological age labels, by contrast, both enriched and random panels fell on the expectation (Fig. 3e, Supplementary Table 2). Meanwhile, median organ selectivity of enriched panels rose from 1.06 (95% CI 0.79–1.85) under AA to 2.00 (95% CI 1.03–3.16) under OS-Morb (Fig. 3f). Sensitivity analyses, including removing NMR metabolomics, adjusting for the shared latent clock component, removing diagnostic markers, leaving one disease out of the label and excluding early events, showed that organ-matched hazard ratios were largely retained and not driven by reverse causation or single diagnostic markers (Extended Data Fig. 1a–e, Supplementary Tables 5 and 6).

To aid clinical interpretation, OS-Morb scores can also be expressed in units of biological age (years) without altering downstream risk rankings. Scaling each clock against the all-cause mortality HR per year of chronological age (HR 1.11 per year, 95% CI 1.10–1.12; HR 1.67 per 5 years, 95% CI 1.62–1.73), analogous to PhenoAge^13^, a 1-SD increase in clock score at age 55 years corresponded to an organ age acceleration ranging from 3.5 years (95% CI 3.0–4.0) for bone to 16.1 years (95% CI 14.9–17.4) for pancreas, with larger year-equivalents observed at younger baseline ages across most organs (Extended Data Fig. 2b, Supplementary Table 4). The immune and intestine clocks, which did not show significant cognate disease associations in the multi-omics sub-cohort, were excluded from year conversion. On average this organ age followed chronological age, with a wider spread than organ age from the AA clock on the same features (Fig. 1c, Extended Data Fig. 2a,c– e); it predicted the organ’s own diseases better than the AA organ age and than PhenoAge, and discriminated as well as the standardized score (Extended Data Fig. 2f–h, Supplementary Table 4). All subsequent analyses used the standardized clock scores directly.

### Outcome-anchored definitions of extreme aging resolve clinical discordance

Defining extreme aging via statistical distribution thresholds creates severe clinical discrepancies across organs. A uniform cut-off of 2 standard deviations (SD)^4^ or 1.5 SD^6^ flags an identical proportion of individuals in every organ by definition (2.27% above 2 SD under a normal distribution; Fig. 4a), yet the resulting 5-year incidence of cognate disease among flagged individuals ranges from 0.2% in the intestine to 43.3% in adipose tissue (Fig. 4b).

**Fig. 4.**
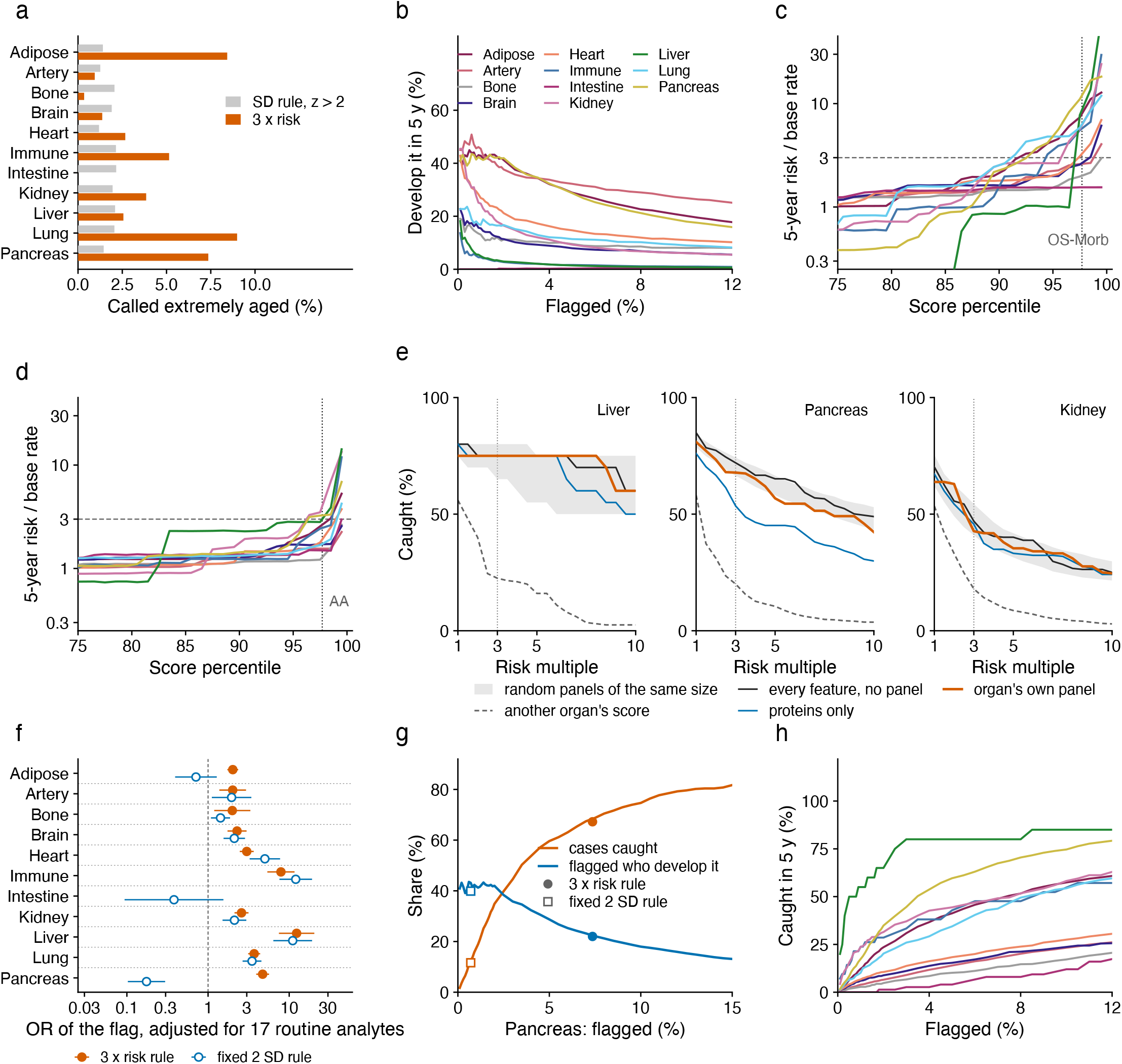
| Two definitions of extreme aging. Multi-omics cohort, n = 22,689; proportions are computed within the evaluable population of each organ. **a**, Proportion of people called extremely aged by organ, one line for a score above 2 SD and one for a 5-year risk reaching three times the base rate. **b**, Proportion flagged and 5-year incidence among the flagged, by organ. **c**, Out-of-fold risk curves by organ (log axis; five-fold out-of-fold isotonic regression); horizontal dashed line, three-fold risk; vertical dotted line, 2 SD. **d**, As **c**, with the training label changed to age. **e**, Proportion of 5-year cases captured at different risk multiples for the liver, pancreas and kidney, comparing the organ’s own panel, the union of panels, proteins only and scores of other organs; shading, 2.5th to 97.5th percentiles of 1,000 random panels; dotted line, three-fold. **f**, Odds ratios of each organ’s flag for the matched disease (log axis), adjusted for demographics and 17 routine tests; lines, 95% CI; dashed line, 1; outcome, new onset of the matched disease during follow-up. **g**, Pancreas: the curves of **b** and **h** side by side; filled point, risk-rule cut-point; open point, SD-rule cut-point. **h**, Proportion flagged and proportion of 5-year cases captured, by organ.

When extreme aging is defined by outcome risk, we convert the clock scores into calibrated 5-year absolute risks of the matched disease and define extreme aging as a risk reaching three times the organ’s baseline incidence (Fig. 4c,d, Extended Data Fig. 3a). Across risk multiples ranging from 1 to 10, case capture generally exceeded the organ-swapped null (Fig. 4e, Extended Data Fig. 3b) and held steady whether models used organ-enriched panels, random panels of equal size, the unconstrained feature pool or proteins alone (Fig. 4e). Furthermore, after adjusting for demographics and 17 routine clinical analytes, risk-anchored flags maintained independent associations with incident disease in all organs (odds ratios above 1), whereas the 2 SD flags did not for the pancreas and adipose tissue (Fig. 4f).

Rather than representing distinct phenomena, score-based and risk-based rules are two cut-points on the same sensitivity–specificity continuum (Fig. 4g). When the SD cut-point is moved to flag the same number of people as the risk rule, the two rules select largely the same people (Fig. 4h, Supplementary Table 7). Anchoring thresholds to absolute risk accommodates baseline epidemiological variation, yielding organ-tailored identification rates that range from 0% in low-incidence organs (intestine) to 9.0% in high-incidence organs (lung) (Fig. 4a).

### Clocks trained on routine laboratory tests transfer robustly across cohorts

To establish translational utility, we assessed whether organ clocks could be derived exclusively from routine laboratory tests, avoiding high-dimensional omics. We systematically stepped down feature inputs from 913 multi-omics features, to 276 clinical biochemistry and NMR measures, to 33 routine laboratory analytes shared by the UK Biobank and NHANES, and finally to a parsimonious panel of 16 standard clinical tests (10 for the JMDC cohort), training and evaluating in 390,087 UK Biobank participants (Fig. 5a).

**Fig. 5.**
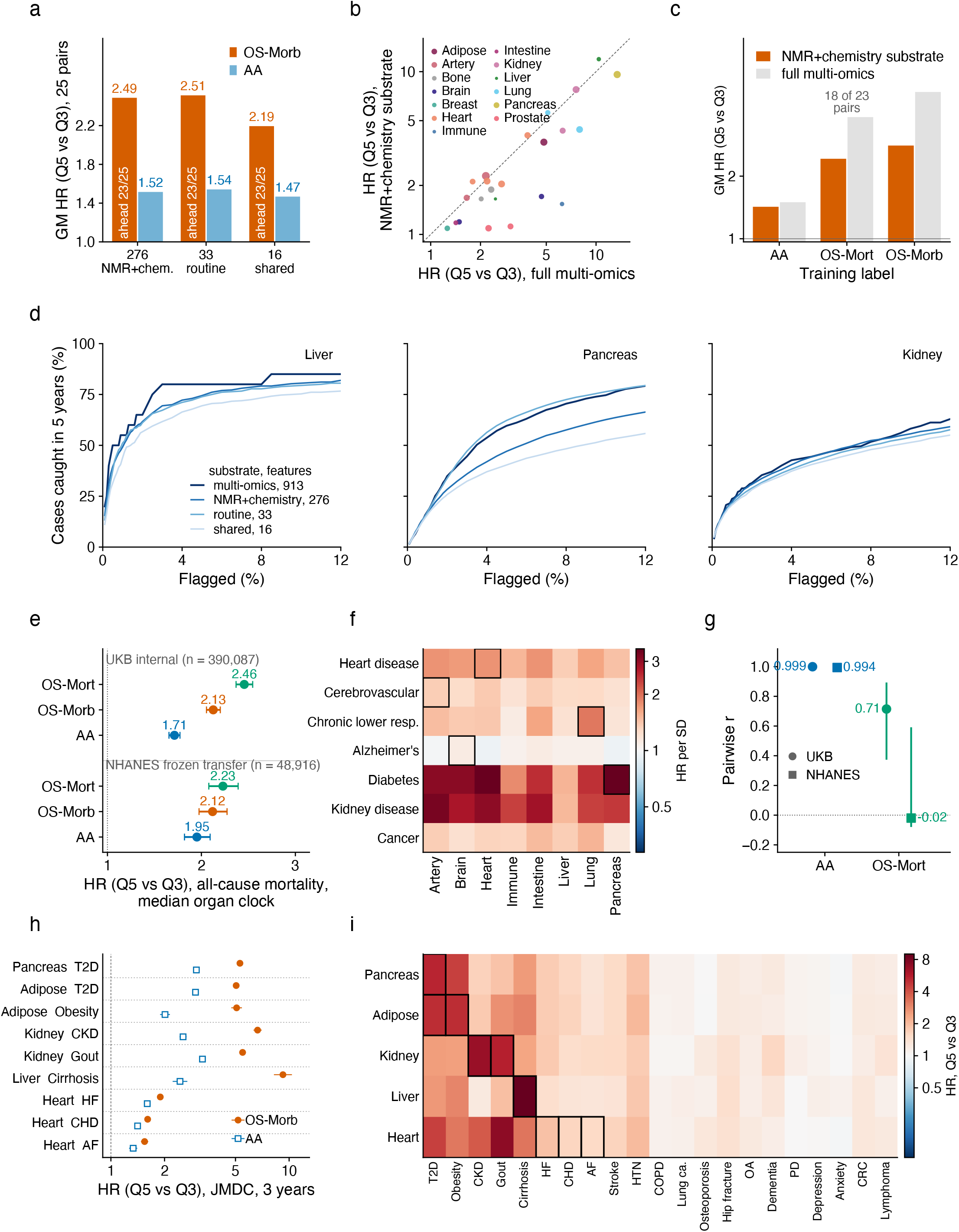
| Organ clocks trained on routine laboratory tests transfer to an independent cohort. **a**, Geometric mean Q5 versus Q3 hazard ratio of OS-Morb against AA trained on the same features on three substrates, with counts of pairs higher and lower; multi-omics cohort, n = 22,689. **b**, Q5 versus Q3 hazard ratios of 23 pairs on NMR + chemistry and on full multi-omics. **c**, Geometric mean hazard ratio on the two substrates under the three training labels, over pairs with readouts on both substrates; under OS-Mort, 5 pairs on the cheap substrate were not fitted because own-organ deaths were fewer than 50, as marked. **d**, Proportion flagged and proportion of 5-year cases captured for the liver, pancreas and kidney on four substrates; multi-omics n = 22,689, other three n = 390,087. **e**, Q5 versus Q3 hazard ratio for all-cause mortality with 95% CI, within the UKB and after frozen transfer to NHANES; each label is represented by the median of its organ clocks. **f**, Hazard ratio per SD of each OS-Mort organ clock for seven NHANES causes of death after frozen transfer (log colour scale); boxes, death from the clock’s own organ. Random-direction null tests are given in Supplementary Table 6. **g**, Median pairwise correlation between organ clocks in two cohorts × two labels; lines, minimum to maximum. **h**, JMDC, n = 3,190,470. Q5 versus Q3 hazard ratios for first onset within 3 years in 9 organ–disease pairs, OS-Morb and AA trained on the same 10 tests; lines, 95% CI. **i**, JMDC: Q5 versus Q3 hazard ratios of 5 OS-Morb organ clocks for 21 diseases over 3 years; boxes, each clock’s own diseases.

Although information loss was observed, predictive performance remained robust. In the 22,689 people with complete data on all four substrates, the geometric mean HR decreased from 3.34 (95% CI 2.52–4.68) for full multi-omics to 2.49 (95% CI 1.84–3.49) with NMR + chemistry (Fig. 5b,c) and 2.08 (95% CI 1.59–2.79) with the 16-test routine panel (Supplementary Table 4). Outcome-anchored case capture curves on the three cheaper substrates showed consistent scaling: the cheaper the substrate, the fewer cases were captured at the same proportion flagged (Fig. 5d, Extended Data Fig. 4a, Supplementary Table 7).

UK Biobank OS-Morb models applied without coefficient calibration to the US NHANES cohort maintained both discrimination and organ selectivity. The all-cause mortality HR (Q5 versus Q3) for the median OS-Morb clocks was 2.12 (95% CI 1.98– 2.27) in NHANES and 2.13 (95% CI 2.05–2.20) in the UK Biobank (Fig. 5e). In NHANES, baseline OS-Morb clocks were associated with cognate self-reported diagnoses, with the prevalence odds ratios of the own-organ clock exceeding the random null for all 8 conditions (Extended Data Fig. 4b). By cause of death, the transferred OS-Mort pancreas clock had an HR per SD of 14.8 for diabetes mortality, compared to an average of 1.4 across non-pancreas clocks (Fig. 5f). Moreover, OS-Mort clocks retrained locally on NHANES mortality data discriminated all-cause death as well as the transferred clocks (C-index 0.856 versus 0.867) but were nearly uncorrelated with one another (median pairwise correlation −0.02; Fig. 5g), and none of them retained attribution to death from its own organ (Supplementary Table 6).

In the Japanese health check-up and claims cohort JMDC, the transferred clocks also predicted first onset of disease of their own organ. We then fixed the coefficients of the 10-test clocks and applied them to 3,190,470 JMDC participants aged 40 to 74 years, with a median follow-up of 3.3 years. For first onset within 3 years, the Q5 versus Q3 hazard ratio was 9.23 (95% CI 8.22–10.37) for the liver clock and cirrhosis, 6.65 (95% CI 6.31–7.01) for the kidney clock and chronic kidney disease, and 5.30 (95% CI 5.19–5.40) for the pancreas clock and type 2 diabetes (Fig. 5h,i, Supplementary Table 6). Among the five transferred organ clocks, the own-organ clock had the highest hazard ratio for each of these three diseases. OS-Morb exceeded AA trained on the same 10 tests in all 9 pairs (Extended Data Fig. 4c). The organ clocks added discrimination beyond age and sex. For first onset within 3 years, the C-index increment ΔC was 0.193 (95% CI 0.185–0.204) for the liver clock and cirrhosis, 0.136 (95% CI 0.132–0.140) for the kidney clock and chronic kidney disease, and 0.121 (95% CI 0.119–0.123) for the pancreas clock and type 2 diabetes. With 3-year risk refitted within JMDC on age, sex and the clock score, predicted incidence agreed with observed incidence across deciles (Extended Data Fig. 4d).

### Organ clocks improve risk stratification beyond current clinical scores

Adding OS-Morb clocks to the current clinical risk score of each organ (SCORE2^21^; QRISK3^22^; QDiabetes^23^; FINDRISC^24^; eGFR^25^; kidney failure risk equation^26^; FIB-4^1^; NAFLD fibrosis score, NFS^27^) improved disease discrimination across organs, most for the liver and least for the heart and artery. Incremental C-index improvements (ΔC) were modest for cardiovascular models (heart clock on SCORE2, ΔC 0.016, 95% CI 0.015–0.018; artery clock similar), but substantial for hepatic assessment (liver clock on FIB-4, ΔC 0.126, 95% CI 0.112–0.139), with the pancreas and lung clocks in between (Fig. 6a). The models maintained robust 10-year risk calibration, similar to that of the score alone (Fig. 6b). Clocks derived from 33 routine laboratory tests showed similar performance, notably increasing discrimination for type 2 diabetes when the pancreas clock was added to QDiabetes (ΔC 0.080, 95% CI 0.078–0.083). Reclassification metrics indicated that gains in pancreas and liver models were driven primarily by up-classifying incident cases, whereas gains in heart and artery models were driven by down-classifying non-cases (Fig. 6c, Supplementary Table 8).

**Fig. 6.**
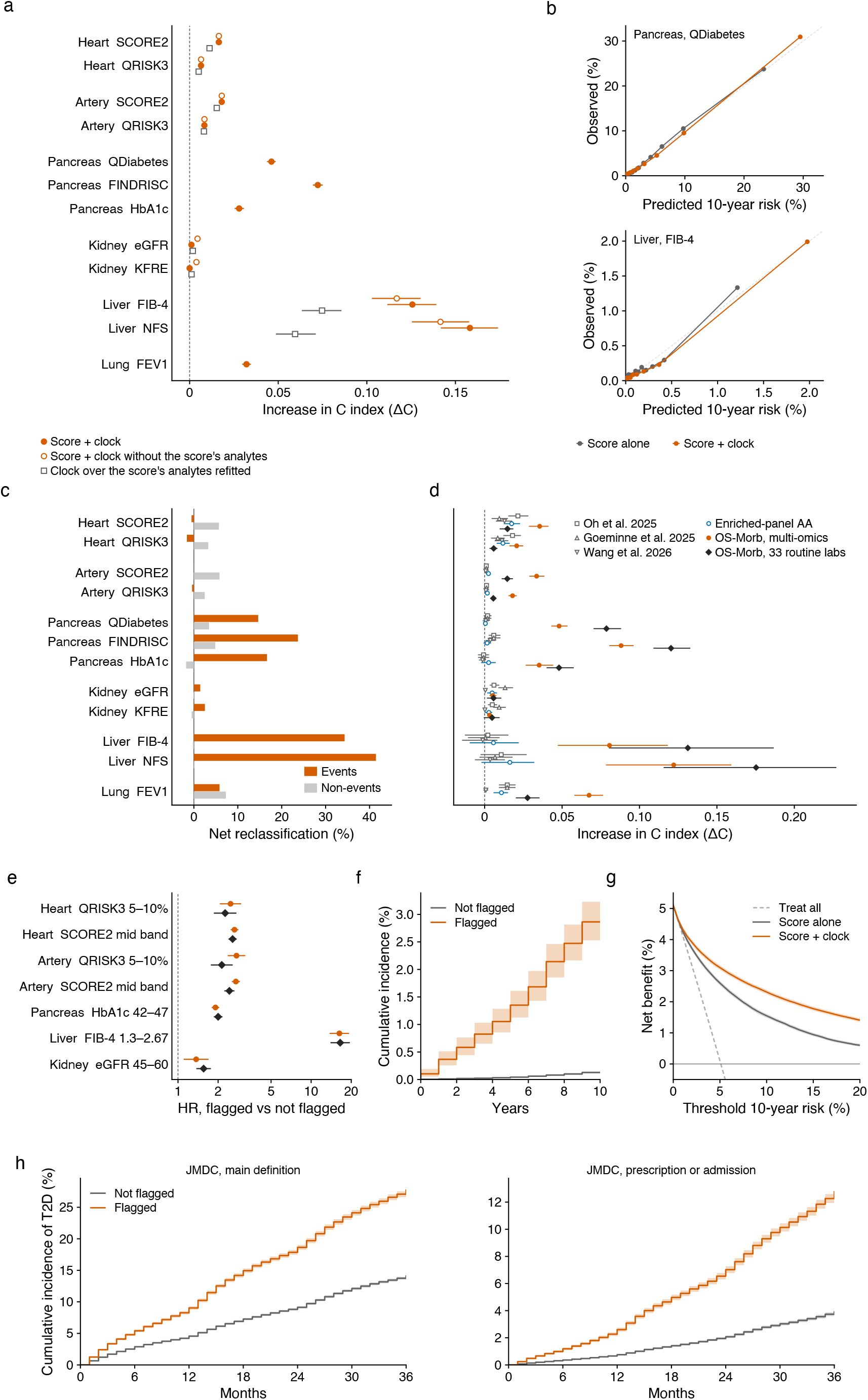
| Increment of organ clocks over clinical risk scores. NMR + chemistry cohort, n = 390,087. **a**, C-index increment after adding the organ clock, for each organ and score. **b**, 10-year risk calibration of score alone and score plus clock for QDiabetes (pancreas) and FIB-4 (liver). **c**, Net reclassification of cases and non-cases after adding the clock. **d**, Multi-omics cohort, n = 22,689. C-index increment on the same score after adding organ age clocks rebuilt from three published designs, the enriched-panel age clocks and the OS-Morb clocks (multi-omics and 33 routine tests), in the same people for each score. **e**, Within the inconclusive range of each score, hazard ratio of people flagged by the organ clock against those not flagged. **f**, 10-year cumulative incidence among people with FIB-4 between 1.3 and 2.67, by liver-clock flag. **g**, Decision curve for QDiabetes (pancreas). **h, External validation in JMDC (Japan), n = 168,680 people with HbA1c 42–47 mmol/mol: cumulative incidence of type 2 diabetes over 3 years in people flagged and not flagged by the pancreas clock (10 routine tests, coefficients trained in the UK Biobank, same flag rule as in e). Left, main outcome definition; right, strict definition requiring a prescription or admission.** Lines and shading, 95% CI.

OS-Morb organ clocks added more information to the scores than current organ age clocks. In the multi-omics cohort, we rebuilt organ age clocks following Oh et al.^20^, Goeminne et al.^5^ and Wang et al.^6^ and, together with the age clocks we trained on enriched panels, added each of them and the OS-Morb clocks in turn to the same score in the same people. In 9 of 12 organ and score combinations, the ΔC of the OS-Morb clock exceeded that of all four age clocks. The ΔC of the heart clock on SCORE2 was 0.036 (95% CI 0.029–0.041), 0.014 (0.008–0.020) higher than that of the heart clock of Oh et al.^20^. On the five pancreas and liver scores, no age clock added more than 0.016, whereas the OS-Morb clock on 33 routine tests added 0.048 to 0.175 (Fig. 6d, Supplementary Table 8).

This added predictive value persisted even after adjusting for the individual biochemical components of the liver scores. FIB-4 and NFS together share AST, ALT and albumin with the liver clock; re-entering and refitting these variables before adding the liver clock still yielded a substantial ΔC of 0.075 (95% CI 0.064–0.085) on FIB-4 and 0.060 (95% CI 0.049–0.071) on NFS (Supplementary Table 8).

Finally, organ-specific clocks provided actionable risk stratification among individuals in clinical “gray zones”, the ranges that guidelines consider inconclusive and in need of further testing. Among 151,832 individuals with borderline FIB-4 scores (1.30–2.67), the liver clock flagged only 5.7% (95% CI 5.6–5.9%), yet this small subset accounted for 52.2% (95% CI 48.4–56.0%) of all incident cirrhosis occurring within the intermediate band. Similarly, among individuals with prediabetic HbA1c (42–47 mmol/mol), those flagged by the pancreas clock had a 5-year diabetes incidence of 23.4% (95% CI 22.1–24.8%) compared to 9.9% (95% CI 9.3–10.5%) in non-flagged individuals (Fig. 6e,f). Decision curve analysis confirmed that integrating the pancreas clock into QDiabetes yielded greater net benefit than the score alone across 10-year risk thresholds from 2% to 20%, with non-overlapping 95% intervals (Fig. 6g). In JMDC, among 168,680 people in the same HbA1c range, 27.7% (95% CI 27.2–28.2%) of those flagged by the pancreas clock under the same rule were diagnosed with type 2 diabetes within 3 years, compared to 14.1% (95% CI 13.8–14.3%) of those not flagged (Fig. 6h, Supplementary Table 6). Some flagged individuals already met diagnostic criteria at baseline without a diagnosis. For example, 27.2% (95% CI 26.7–27.8%) of individuals flagged by the pancreas clock already had HbA1c of 48 mmol/mol or above, compared to 0.8% (95% CI 0.8–0.9%) of those who were not flagged. For the kidney clock, 36.2% (95% CI 35.5–36.8%) of those flagged already had eGFR below 60 mL/min/1.73 m² or UACR of 30 mg/g or above, compared to 3.9% (95% CI 3.9–4.0%) of those who were not flagged.

### Clinical endpoints decouple the shared genetic architecture of organ clocks

Genome-wide association studies (GWAS) demonstrated that switching the training objective to organ disease markedly reduced shared genetic correlation across organ clocks (Extended Data Fig. 5a). In the multi-omics cohort, AA clocks built from organ-enriched panels, which is current practice, exhibited an average pairwise genetic correlation (rg) of 0.58 (95% CI 0.47–0.70); combining assigned features with the OS-Morb objective lowered it to 0.35 (95% CI 0.26–0.47; Supplementary Table 9). This decoupling was striking at the locus level. In the NMR + biochemistry cohort (n = 390,087), 35.5% of the 868 genome-wide significant OS-Morb loci mapped to a single organ clock, compared to only 12.3% of the 284 loci for the AA control on the same features (Extended Data Fig. 5b,c, Supplementary Table 10). Furthermore, OS-Morb clocks captured genuine organ-specific genetic liability. Across 30 external disease GWAS, OS-Morb clocks had higher genetic correlation with cognate than with non-cognate diseases (Δrg +0.10, 95% CI 0.00– 0.21), whereas the AA control on the same features showed no excess (Δrg −0.01, 95% CI −0.06 to 0.04; Extended Data Fig. 5d,e, Extended Data Fig. 6).

## Discussion

This study shows that organ specificity in blood-based aging clocks is governed primarily by the training label rather than by a priori tissue-enriched feature selection. By replacing chronological age with the first onset of organ-specific disease (OS-Morb), organ clocks can be built directly from standard routine laboratory tests, without proteomics or metabolomics. Trained once in the UK Biobank and deployed with frozen coefficients, these routine-test organ clocks kept organ selectivity in the US NHANES cohort, predicted first onset of disease of their own organ among 3.19 million people in the Japanese JMDC cohort, and added more discrimination over established clinical risk scores than current proteomic organ age clocks. More importantly, the new approach enhances the ability to stratify individuals/phenotypes relative to traditional models.

The most immediate application is multi-organ risk profiling from a single routine blood draw. Because the input tests are already measured in annual primary-care and occupational health checks, an organ-resolved risk panel needs no specialized assay platform and has close to zero marginal cost. In clinical decisions, routine-test organ clocks are most useful for people whose conventional single-organ scores fall in an inconclusive range, where they can help triage confirmatory testing, prioritize limited preventive resources and detect unrecognized disease at baseline. For example, when FIB-4 falls in the indeterminate range (1.30–2.67), current hepatology guidelines recommend second-line transient elastography or a 1-year lifestyle intervention followed by repeat testing^2^. Referring only the 5.7% of people in this range flagged by the routine-test liver clock would capture 52.2% of subsequent cirrhosis cases in the range while sending fewer than one in ten people on to further testing. Similarly, where places in diabetes prevention programmes are limited and eligibility is ranked by HbA1c alone^3^, the pancreas clock provides an additional, outcome-calibrated ranking within the prediabetic range (42–47 mmol/mol) that was replicated in both the UK Biobank and JMDC. Moreover, because more than a quarter of pancreas-flagged and a third of kidney-flagged people without a prior diagnosis already met biochemical criteria for diabetes or chronic kidney disease at baseline, routine-test clocks can surface missed diagnoses from existing test results.

For population screening, anchoring extreme-aging flags to multiples of baseline disease risk addresses a limitation of fixed statistical cut-offs. A uniform parametric threshold (such as 2 SD or 1.5 SD) flags the same fraction of the population in every organ by construction, although 5-year disease incidence varies by two orders of magnitude across organ systems. Defining extreme aging as a risk multiple (such as three times the baseline incidence) instead aligns the share flagged with disease burden and ties every flag to an explicit absolute risk. Clinicians and health systems can therefore choose operating thresholds that reflect the trade-off between missed cases and false alarms^28,29^. Our external validations suggest a two-step deployment: coefficients trained in a deeply phenotyped cohort with long follow-up for disease onset are frozen and transferred, and only the absolute risk threshold is reset from local incidence. Where a cohort records only deaths, as NHANES does, clocks retrained on local cause-specific deaths kept the discrimination of the transferred clocks, but none pointed to death from its own organ, which favours freezing clocks trained where disease onset is followed.

Conceptually, defining organ aging through organ-specific morbidity extends the approach of second-generation epigenetic clocks from the whole organism to individual organs. First-generation clocks trained on chronological age capture time-dependent molecular drift^30^, much of which reflects systemic rather than organ-specific change, whereas PhenoAge and GrimAge define biological aging through its consequences for disease and death^13,14^. OS-Morb carries this idea to the organ and, like PhenoAge, can state organ aging in years. This explains why unconstrained OS-Morb models up-weighted proteins enriched in their own organ, whereas unconstrained chronological-age models converged on a single systemic signature. Our comparison with a pan-disease clock (AC-Morb) separates two steps: switching the label from chronological age to disease provides most of the gain in risk discrimination, and restricting the disease composite to the organ directs each clock toward its own pathology. The genetic analyses agree: switching from chronological age to OS-Morb lowered cross-organ genetic correlation by about 40%, from 0.58 to 0.35, and nearly tripled the share of loci reaching genome-wide significance for only one organ clock. Once organ identity is encoded in the label, tissue-assigned protein selection is no longer needed, and routine clinical chemistry can serve as the substrate.

Our null-model audits also put findings from earlier organ and system clocks in context. The low inter-clock correlations, disease associations and large shares of people with an extremely aged organ reported for chronological-age proteomic clocks^4–6,20^ are reproducible observations; however, size-matched random panels and the equicorrelated Gaussian model show that these properties are expected from sparse feature partitioning and the geometry of regression residuals, without organ-specific biology. Multi-system clocks built from routine biomarkers grouped a priori and trained on chronological age^31,32^ likewise have low inter-clock correlations, but lower hazard ratios for disease of their own organ than disease-trained clocks. In head-to-head comparisons in the same participants, chronological-age clocks rebuilt from three published proteomic schemes added less discrimination to established clinical risk scores than OS-Morb clocks, and almost none to the liver and pancreas scores. Single-disease blood risk scores, for their part, are not organized by organ^33–35^.

Several limitations point to future work. First, the models were trained in the UK Biobank, whose participants are mostly of European ancestry and healthier than the general population^36^. Frozen transfer to the multi-ethnic US NHANES cohort and the Japanese JMDC cohort tested performance in other populations but not across ancestries; JMDC also had only 10 of the routine tests, a median follow-up of 3.3 years and no adults aged 75 years or older. Validation in further cohorts with more ancestries and longer follow-up is warranted. Second, mapping diagnoses to organs relies on diagnosis coding and on grouping decisions. Because the brain composite was dominated numerically by depression and anxiety, the brain clock was insensitive to rarer neurodegenerative disorders such as Parkinson’s disease; dividing organ labels by mechanism (for example, separating neurodegenerative, vascular and psychiatric brain endpoints) should sharpen resolution. Third, the clocks were derived from baseline blood measurements at a single time point and cannot separate baseline risk from the pace of organ decline; applying the frozen clocks to repeated annual health-check blood draws will allow individual organ-aging trajectories to be tracked. Fourth, each clock was trained and evaluated for disease of one organ at a time; how risks combine across organs in people with multimorbidity was not modelled, and joint multi-organ models are a natural next step.

In summary, organ aging can be defined and measured through organ-specific disease risk. With organ identity placed in the training label rather than in tissue-assigned features, organ clocks can be built from standard laboratory tests, frozen and transferred across populations. This offers a practical, scalable route to organ-resolved risk stratification in routine healthcare.

## Methods

### Study population and data

All discovery analyses used the UK Biobank^15^. Four nested cohorts were used. Every number in the paper comes from one of them. Cohort A, the Olink proteomic cohort (n = 52,614), was used for the audit of published organ clocks and for the protein-panel null models. Cohort B, the multi-omics cohort (n = 22,689), comprised participants with Olink proteomics, NMR metabolomics, clinical chemistry and hormones, an age and a follow-up timeline. It supplied the main hazard ratios and the comparison of feature provision and training labels. Cohort B′ comprised the members of cohort B who could be scored on both a cheap substrate and the multi-omics substrate (13,015 to 28,594 per disease, depending on the risk set). Cohort C, the NMR and chemistry cohort (n = 390,087; 211,245 women and 178,842 men), comprised all participants with complete scores on that substrate. Organ-specific death phenotypes covered all 487,713 participants and were intersected with each cohort. The UK Biobank data were used under Application Number 66813.

Two external cohorts were used for transfer. The US National Health and Nutrition Examination Survey (NHANES, ten cycles 1999 to 2018)^16^ provided adults aged 20 years or older who completed the examination, with mortality linkage to the National Death Index (public release 2019, median follow-up 9.2 years). The Japan Medical Data Center (JMDC) claims and health check-up database^17^ was analysed under the approved project “Health trajectories and treatment effects across adulthood”. Individual-level JMDC data did not leave the JMDC computing environment; only aggregate results were exported, with counts rounded to the nearest 10 and cells with fewer than 10 people suppressed.

UK Biobank has approval from the North West Multi-centre Research Ethics Committee (REC reference 21/NW/0157), and all participants gave written informed consent. NHANES protocols were approved by the National Center for Health Statistics Research Ethics Review Board, and participants gave written informed consent. JMDC data are anonymously processed information under the Japanese Act on the Protection of Personal Information and were used under licence from JMDC Inc.; research using only such data falls outside the scope of the Japanese Ethical Guidelines for Medical and Biological Research Involving Human Subjects, and individual consent was not required. FinnGen data were used only as publicly available summary statistics.

### Feature substrates

The NMR and chemistry substrate comprised all 251 Nightingale NMR measures at the baseline visit (including derived ratios, without pre-filtering), 25 routine clinical chemistry tests and sex, 276 features in total^37^. On this substrate every organ clock received the same feature matrix, so clocks differed only in their training label. NMR measures and hormones were log(x + 1) transformed; Olink values were used as NPX.

The multi-omics substrate combined Olink proteins^38^, NMR measures, clinical chemistry and hormones. Feature assignment to organs applied to proteins only. Each organ received the Olink proteins annotated as tissue-enriched or group-enriched for that tissue in the Human Protein Atlas^18^; GTEx tissue enrichment^39^ was used only for post hoc validation. The assigned panels were expressed by the expected parenchymal cells in the Tabula Sapiens single-cell atlas^40,41^; for example, 80 of the 131 liver proteins were expressed and 62 mapped to hepatocytes (Supplementary Table 11). In the assigned arm, each organ received its protein panel (12 to 187 proteins) plus 233 NMR measures and 24 routine clinical measures shared by all organs (the nine PhenoAge inputs and 15 organ-related tests such as ALT and bilirubin for the liver). In the unassigned arm, every organ received the union of all panels: 656 proteins, 233 NMR measures and 24 clinical measures, 913 features in total. Assigning the 15 organ-related clinical tests to organs as well was reported as a sensitivity analysis (Supplementary Table 6).

Two routine-test substrates were built on cohort C. The 33-test panel contained the tests measured both at the UK Biobank baseline and in all ten NHANES cycles: 16 biochemistry tests (albumin, alkaline phosphatase, ALT, AST, GGT, urea, creatinine, glucose, urate, total bilirubin, triglycerides, total cholesterol, calcium, phosphate, total protein and HbA1c), 15 blood-count measures, HDL cholesterol and CRP, plus sex. The 16-test panel was the most basic subset of these (albumin, alkaline phosphatase, ALT, AST, urea, creatinine, glucose, urate, total bilirubin, triglycerides, white cell count, MCV, RDW, lymphocyte percentage, HDL cholesterol and CRP) plus sex. For JMDC, the UK Biobank clocks were retrained on the 10 tests shared with the Japanese health check (ALT, AST, GGT, creatinine, urate, glucose, HbA1c, HDL cholesterol, LDL cholesterol and triglycerides) plus sex.

For the audit of published clocks in cohort A, protein panels of 9 to 227 Olink proteins per organ were taken from published organ assignments. The SomaScan panels of Oh et al.^4^ were mapped to Olink by gene symbol, covering 10 organs with 3 to 95 proteins each.

### Training labels

Four labels were used with an otherwise identical pipeline. Chronological age gave the conventional age clock (AA). Organ-specific morbidity (OS-Morb) was first onset, during follow-up, of any disease in the organ’s own disease set (first ICD-10 diagnosis in linked records); participants with any of these diseases at baseline were excluded from training. All-cause morbidity (AC-Morb) was first onset of any of the 22 diseases, one clock per organ on that organ’s assigned panel. This label separates the effect of using a disease label from the effect of matching the label to the organ (Supplementary Table 4). Organ-specific mortality (OS-Mort) was death with an underlying cause (UK Biobank field 40001, one ICD-10 code per death) in a fixed set of three-character code blocks assigned to the organ, built from the same disease table plus the organ’s cancers and organ failure codes (Supplementary Table 1). Each death was assigned to at most one organ, deaths from other causes were censored at the date of death, and deaths whose underlying cause fell in no organ block entered no organ label. OS-Mort was not trained for organs with fewer than 100 deaths. Breast and prostate clocks were trained and evaluated within one sex.

Incidence and death labels were coded 0 or 1 and fitted with the same elastic-net regression as age, a penalized linear probability model. Cox models were used only for evaluation. Three disease tables were kept separate. The composite membership table defined the training labels (22 diseases; Supplementary Table 1). The pair table defined the 25 organ–disease pairs used to evaluate hazard ratios, drawn from 11 organs and 22 diseases, with type 2 diabetes, coronary heart disease and acute myocardial infarction each shared by two organs. The transfer table contained 30 single diseases, each with its own clock.

### Model fitting

The same seven steps were used for every organ and label (Fig. 1b): median imputation, standardization, choice of the elastic-net penalty by cross-validation, outer five-fold out-of-fold prediction so that no participant was scored by a model that had seen them, linear residualization of the score on chronological age, Blom rank-normal transformation, and Cox evaluation^42–44^. The Blom transformation, z = Φ⁻¹((r − 0.375)/(n + 0.25)), was applied once to the whole scored population and not repeated within risk sets. The OS-Morb training outcome and the evaluation outcome come from the same records. The outer out-of-fold split and the AC-Morb comparison guard against this circularity.

### Survival analysis

Cox proportional hazards models were fitted without a penalty (lifelines CoxPHFitter)^45,46^. Some clinical-score models did not converge at first; for these, the Newton step was reduced to 0.5 and then 0.1. Models were adjusted for chronological age and sex (sex was omitted for sex-specific organs), and all compared scores (OS-Morb, AA, PhenoAge and AC-Morb) used the same risk set and adjustment. PhenoAge was computed as in Levine et al.^13^. Hazard ratios compare the top with the middle quintile of score (Q5 versus Q3). Quintiles were formed by rank within each pair’s risk set, and a Cox model was fitted to an indicator of Q5 against Q3. The middle quintile was used as reference because the lowest quintile often contains reverse pathology. Per-SD models were used only for null-model direction counts and for the NHANES organ-matching contrast. Time ran from baseline to first diagnosis, and people without an event were censored at the end of follow-up. People with the disease at baseline were excluded. Pairs with fewer than 20 events were not reported. For organ-specific death, deaths from other causes were censored at the date of death (cause-specific hazards)^47^; subdistribution hazards were not estimated^48^. Models were not adjusted for BMI, smoking or deprivation. Instead, diagnostic markers were removed to address feature-level circularity, and landmark analyses addressed reverse causation.

In the landmark analyses^49^, people with an event or censoring within L years (L = 2 or 5) were removed and time was reset to time − L; everything else was unchanged. Diagnostic markers that enter the outcome definition were removed in a first tier, and adjacent markers (lipids for cardiovascular outcomes, white-cell indices for lymphoma, smoking history for the lung) in a second tier. In the substrate test, all NMR measures were removed from the NMR and chemistry panel.

Every quantity estimated from a sample is given with a 95% interval. Cox and logistic coefficients use Wald intervals; proportions use Wilson score intervals^50^; aggregates across organs or pairs (geometric means, medians and means) use cluster bootstraps over organs or pairs (2,000 resamples) with the 2.5th and 97.5th percentiles as limits; correlations use Fisher z intervals^51^; comparisons with null models also give the 2.5th to 97.5th percentiles of the null distribution^52^. Counts, panel sizes, preset thresholds and sample sizes carry no interval. The 25 pairs come from 11 organs and 22 diseases and are not independent. Win counts were therefore tested with an organ-level cluster bootstrap (11 organs resampled with replacement, 10,000 times), which gives a 95% interval for the win share and P(share ≤ 0.5). The share was also reported with each disease counted once.

Discrimination was assessed as the gain in Harrell’s C over two base models, age and sex, and age, sex, BMI, smoking and systolic blood pressure^53^. We also report the ratio of event rates between the top and bottom score deciles and the calibration slope of the linear predictor.

### Organ age in years

Following the way PhenoAge converts mortality risk into years^13^, OS-Morb scores of the main clocks (cohort B, organ-assigned panels, out-of-fold) were converted into an organ age:

organ age = chronological age + β(age) × z / γ.

Here z is the age-residualized, Blom-transformed and standardized score; β(age) = b0 + b1 × (age − 55), with age truncated to 40 to 70 years, where b0 and b1 come from a Cox model of first onset of the organ’s own composite on z, the product of z and age, age and sex; and γ is the age coefficient of a Cox model of all-cause death on age and sex in the same cohort (HR 1.11 per year, 95% CI 1.10–1.12; 2,003 deaths). All organs share this denominator, so one year means the rise in mortality hazard brought by one year of age, the unit of PhenoAge. Using each disease’s own age slope instead would inflate the years for diseases that rise slowly with age, such as obesity and depression. The 95% interval of years per SD was obtained by the delta method. No organ age was defined when the lower 95% limit of the clock’s per-SD hazard ratio for its own diseases was 1 or below (immune system and intestine). For comparison with conventional measures, the OS-Morb organ age, organ age from the AA clock on the same organ-assigned features (chronological age plus its out-of-fold age gap) and PhenoAge were each used alone as the predictor of first onset of the organ’s own diseases. The standardized score and the organ age gap were also compared within one Cox model adjusted for age and sex. Harrell’s C was compared with paired differences over 200 bootstrap resamples of people. For display, organ age was plotted against chronological age with the mean and the 2.5th and 97.5th percentiles in each year of age. Results are in Fig. 1c, Extended Data Fig. 2 and Supplementary Table 4.

### Audit of published organ clocks and null models

The expected proportion of people with at least one organ above +2 SD among K clocks with common pairwise correlation r was computed from an equicorrelated multivariate normal model: conditional on a common factor z₀, the age gaps are independent N(√r·z₀, 1 − r), and the probability is 1 − E[Φ((2 − √r·z₀)/√(1 − r))^K], integrated numerically over z₀^54^. With the correlation matrix of the 11 organ-assigned AA clocks of cohort B, a simulation of 400,000 people gave 11.4% against 11.5% for the equicorrelated approximation. Published constructions were placed at their own K and r: our enriched and random panels at K = 12; the Oh scheme rerun in the UK Biobank at K = 10; Oh et al.^4^ at K = 13, mean r 0.29 and a reported 23%; and Wang et al.^6^ at K = 11, mean r 0.16, threshold 1.5 SD and a reported 40% (Supplementary Table 2). For Fig. 3e, the observed proportion minus this expectation was computed for enriched panels and for 1,000 random panels, with 95% intervals from 1,000 bootstrap resamples of people.

Three published protein-selection schemes were rerun with age labels in cohort A following each original pipeline, with five-fold out-of-fold predictions. For Oh et al.^20^, the native Olink organ panels were retrained with LASSO (10 organs). For Goeminne et al.^5^, the organ age models of their Table S1A were retrained with their elastic-net grid after kNN imputation (k = 10; 10 organs). For Wang et al.^6^, the proteins selected by Boruta^55^ in their Supplementary Table 4 were retrained with LightGBM and Optuna over the original search space (150 trials; 9 organs). Age gaps were residualized on age and z-standardized within sex. Out-of-fold age correlations differed from the published values by at most 0.035 (Supplementary Table 2). Each scheme was compared with 1,000 random panels of the same sizes, retrained with the same pipeline and hyperparameters, and with an organ-swap null (1,000 derangements in which the clock of organ j is used for organ i). The all-cause-death organ models of Goeminne et al.^5^ (coefficients from their Table S1C) were compared with our organ-specific death clocks on the geometric mean hazard ratio for own-organ against other-organ death, with 1,000 random panels as null (Supplementary Table 2). System clocks with systems grouped a priori and an age label^31,32^ were rebuilt on cohort C with the Klemera–Doubal BA_E estimator and, as support, with sex-specific support vector regression, against 1,000 random groupings with the same numbers of systems and tests.

Random-panel nulls used panels of the same size drawn at random and passed through the identical pipeline (same cohort, estimator, folds, calibration and evaluation). Extreme proportions, cross-organ correlations, selectivity and disease prediction used 1,000 panels. Pairs whose permutation P reached the floor of 1/1,001 were extended to 10,000. Empirical P values are one-sided: the proportion of random panels at or above the observed value. Equivalence of the extreme proportion with the random-panel null was tested with two one-sided tests at a prespecified margin of 1 percentage point^56^.

To see which proteins each label selects (Fig. 3b), the two unassigned clocks were fitted once on all trainable people, and for each clock the 50 proteins with the largest absolute coefficients were taken. Their share of the clock’s own HPA organ was divided by that organ’s share of the 656 proteins; the median over the 11 clocks is reported with a 95% interval from 10,000 bootstrap resamples of clocks (Supplementary Table 3).

In the random-composite control, the sibling diseases of an OS-Morb composite were replaced by non-own-organ diseases matched one by one on event count, keeping the target disease and the composite size. All allowed combinations were enumerated (6 to 220 per target disease, 1,613 in total), giving an exact null distribution; P = (number of combinations with a hazard ratio at or above the real composite + 1)/(number of combinations + 1). For the ageotype audit, gap statistics for k = 2 to 8 were computed within sex against an elliptical Gaussian null, and the hit rate of the oldest-organ assignment was compared with a within-sex permutation null^57^.

### Outcome-anchored extreme aging

For each organ, the evaluable population comprised people without the organ’s diseases at baseline who were followed for 5 years or had the disease within 5 years. The organ score was mapped to 5-year risk of the matched disease by monotone isotonic regression with five-fold out-of-fold estimates^58^. A person was flagged when this risk reached a multiple of the organ’s 5-year base rate. The main reading used three times the base rate; multiples from 1 to 10 were scanned. Capture was the share of all 5-year cases in the evaluable population that were flagged, and the positive predictive value was the 5-year incidence among the flagged. The organ-swap null used the scores of the other 10 organs under the same label for the same disease and reported their mean; the stricter 97.5th percentile is in Supplementary Table 6. Cut-points are given as the top percentage of scores, which equals the upper-tail probability of the corresponding z. Intervals came from bootstrap resamples of people (200 in cohort B and 50 in cohort C), refitting the mapping each time. The reference rule was the published 2 SD rule (the organ is the person’s highest and z > 2)^4^, computed in the same evaluable population; Wang et al.^6^ used 1.5 SD. In the equal-budget comparison, the SD threshold was moved to flag as many people as the risk rule, and the risk rule was trimmed to the number flagged by the 2 SD rule. In the random-panel control for flags, each organ’s protein panel was replaced by 1,000 random panels of equal size from the shared protein pool. The 24 shared clinical tests, the label, estimator, folds and risk anchoring were kept.

Odds ratios of the flag for new onset of the matched disease were estimated by logistic regression adjusted for age, sex, BMI, systolic blood pressure, Townsend deprivation, smoking and 17 routine tests (the 16 deployment tests plus HbA1c). The value of three times the base rate was examined with decision curves^29^: when missing a case costs b and a false alarm costs c, action is warranted above a predicted risk pt = c/(b + c), with net benefit TP/N − (FP/N) × pt/(1 − pt). For each organ, the cut-point that maximized net benefit was found at several pt and expressed as a multiple of the base rate. The behaviour of the cut-point across subgroups with different base rates (sex, age band and region of assessment centre) is given in Supplementary Table 7.

### Routine tests and external cohorts

For the same-risk-set comparison of cheap and multi-omics substrates (Fig. 5b,c), cheap clocks were retrained within cohort B′ on the shared NMR and chemistry measures (275 features without proteins) or on the 33-test panel, with the same pipeline, and compared with the organ-assigned multi-omics clocks on 23 pairs; the risk set for each disease comprised people with both scores and without the disease at baseline.

For NHANES, only the UK Biobank coefficients were transferred. Each test was standardized within NHANES (missing values set to 0, the cohort mean), and the score was residualized on age and Blom-transformed within NHANES. The transfer therefore tested discrimination; scale and calibration were not transferred. CRP was not measured in 2011 to 2014 and was set to the mean in those cycles. The main analysis used adults aged 20 or older without survey weights. The question was the relative performance of the three labels in the same people. Weighted Cox models (examination weights pooled over ten cycles, robust standard errors), the 40 to 75 year subset and the non-Hispanic white subset were sensitivity analyses (Supplementary Table 6). NHANES has death follow-up only, with ten underlying-cause groups^59^, so organ matching was possible for the heart (heart disease), lung (chronic lower respiratory disease), artery (cerebrovascular disease), brain (Alzheimer’s disease) and pancreas (diabetes). People with self-reported diagnosed conditions were removed organ by organ in a further sensitivity analysis. Random-direction nulls were recomputed with 1,000 directions in each setting.

For JMDC, the OS-Morb and AA clocks were retrained in cohort C on the 10 shared tests. The coefficients of the five organs whose own-organ signal was best kept in the UK Biobank (pancreas, kidney, liver, heart and adipose tissue) were then frozen and transferred with the same steps as for NHANES (winsorization at the 0.1st and 99.9th percentiles, within-cohort z-scores, age residualization and Blom transformation). Fasting status differed (random glucose in the UK Biobank, about 93% fasting in JMDC); both were standardized within cohort. The cohort comprised each person’s first health check at age 40 to 74 with all 10 tests and at least 12 months of enrolment before baseline (n = 3,190,470; 60% men). Outcomes were first onset within 3 years, requiring a confirmed (not suspected) ICD-10 diagnosis plus at least one supporting record: a confirmed diagnosis in another month, a matching prescription, or a hospital admission with that diagnosis. People with the disease during the look-back period were excluded for that disease. A strict definition accepting only prescriptions or admissions as support was a sensitivity analysis. Follow-up ended at the outcome, disenrolment, age 75 or the data cut-off. Q5 versus Q3 hazard ratios were estimated from unpenalized Cox models adjusted for age and sex. Discrimination was the gain in Harrell’s C over age and sex for first onset within 3 years, with 95% intervals from 200 bootstrap resamples refitting both models. Calibration was assessed out of fold. In each of five folds, a Cox model of age, sex and the clock score (restricted cubic spline, knots at the 5th, 35th, 65th and 95th percentiles) was fitted on the other four folds, and 3-year risk was computed from the Breslow baseline hazard. Predicted and observed (1 − Kaplan– Meier) incidence were compared by decile. Temporal validation fitted on baselines up to 2015 and tested on 2016 onwards. Sensitivity analyses included a 24-month look-back, exclusion of events in the first 12 months, allowing up to two missing tests with mean imputation, and stratification by sex, age band and insurer type (Supplementary Table 6).

### Increment over clinical risk scores

In cohort C, each organ’s OS-Morb clock on the NMR and chemistry substrate was compared with the organ’s current clinical risk score, with the 33-test clock as a sensitivity analysis. Clock scores were five-fold out of fold, and score models were fitted and evaluated on the same folds. Inputs not in the main extract (family history, waist circumference, self-reported ethnicity, FEV1, urine albumin and creatinine, self-reported conditions and clinical total cholesterol) were linked through the identifier map; all 390,087 participants were matched. The scores were SCORE2^21^ and QRISK3^22^ for the heart and artery; FINDRISC^24^ and QDiabetes (model A, without glucose or HbA1c)^23^ for the pancreas, with HbA1c alone as a further comparator; eGFR from creatinine and cystatin C^25^ and, where the urine albumin-to-creatinine ratio was available, the four-variable kidney failure risk equation^26^ for the kidney; FIB-4^1^ and the NAFLD fibrosis score^27^ for the liver; and FEV1 for the lung. SCORE2 was recalibrated to the low-risk region. Outcomes were the organ’s OS-Morb composite for the heart and artery, type 2 diabetes for the pancreas, chronic kidney disease for the kidney, cirrhosis for the liver, and COPD and lung cancer for the lung.

For each organ, four Cox models were fitted: the score alone; the score plus the clock; the score plus a clock retrained without the score’s input tests; and the score’s raw inputs entered one by one plus the clock. The main measure was the gain in Harrell’s C, with 95% intervals from 2,000 bootstrap resamples. Whether an added term carried information was judged by the likelihood-ratio test of nested models^60^ rather than by a DeLong test on the gain in C^61^. Secondary measures were the categorical net reclassification index reported separately for events and non-events at each score’s usual 10-year thresholds^62,63^, which can be positive for uninformative markers under misspecification^64^, calibration slopes, predicted against observed 10-year risk by decile^65^, and 5- and 10-year AUC. Reporting followed TRIPOD+AI^66^, and the framework for incremental value followed Hlatky et al.^67^.

Within each score’s inconclusive range (FIB-4^2^ 1.30 to 2.67; HbA1c^3^ 42 to 47 mmol/mol; QRISK3 10-year risk 5% to 10%; the middle SCORE2 band of the ESC; eGFR 45 to 60), flagged and unflagged people were compared with Cox models adjusted for the score, and 10-year cumulative incidence was estimated with the Aalen–Johansen estimator with death as a competing event. The share of flagged people who already met diagnostic criteria at baseline without a diagnosis (HbA1c at least 48 mmol/mol for the pancreas; eGFR below 60 mL/min/1.73 m² or albumin-to-creatinine ratio at least 3 mg/mmol for the kidney) was given with Wilson intervals. Decision curves compared the score alone with the score plus the clock, with intervals from 2,000 bootstrap resamples.

The head-to-head comparison with current organ clocks (Fig. 6d, Supplementary Table 8) used cohort B. Organ age clocks rebuilt following Oh et al.^20^, Goeminne et al.^5^ and Wang et al.^6^ and our enriched-panel AA clocks (out-of-fold predicted age, residualized on age within the Olink cohort) were compared with the multi-omics, 276-test and 33-test OS-Morb clocks, all added to the same score in the same people, with intervals from 200 bootstrap resamples shared across clocks.

### Genetic analyses

Clock scores on the NMR and chemistry substrate (age label and OS-Morb label, 13 organs) were out of fold, residualized on age and Blom-transformed, because the tails of a linear probability model would otherwise distort association tests. Genome-wide association used PLINK 2.0 (linear regression)^68^ on hard calls from the UK Biobank imputed genotypes^69^, with variants filtered to minor allele frequency at least 0.01 and minor allele count at least 50 (8,187,967 variants), adjusting for age, sex and 10 genetic principal components. The sample was the intersection of cohort C with complete genotypes and covariates (n = 389,328; per-variant n 350,192 to 389,328). This scan was not restricted by ancestry or relatedness. The free LD score regression intercept absorbed residual structure, and a downsampled control with relatedness filtering reproduced the main genetic correlations. For the multi-omics clocks, the same protocol was applied to a 29,978-person genotyped subset after removing relatives up to second degree with KING (cutoff 0.0884)^70^, giving a median per-variant n of 22,407. The downsampled control (n = 22,639) fixed sample size, population and relatedness filtering at the level of the multi-omics cohort.

Genome-wide significance was P < 5 × 10⁻⁸. Within one scan, significant variants were clustered by distance (neighbours within 500 kb joined); across scans, loci were matched by overlap of ±500 kb intervals. Locus counts on equal samples were checked in two further independent subsets of 24,000 people.

Genetic correlations and heritabilities used LD score regression^71,72^ with HapMap3 variants and the European reference panel. Bivariate intercepts were estimated freely, which absorbs the covariance from complete sample overlap among clocks of the same family. For each combination of substrate and label, the 78 pairs among 13 clocks share their clocks and were not treated as independent. A sensitivity analysis removed six regions with large shared effects (GCKR, SLC2A9, LPA, FADS1/2, APOE and CST3; ±500 kb each). Disease summary statistics for 30 diseases came from FinnGen release 10 (Supplementary Table 12)^73^, and MRI organ age gaps from Cao et al.^74^.

### Statistical reporting

The main readings are effect sizes with 95% intervals rather than counts of significant tests. Counts of organs exceeding a null model were reported at a Benjamini–Hochberg false discovery rate of 5% over the organs tested^75^. Empirical permutation P values are one-sided with a resolution set by the number of draws; values at the floor are marked as “≤”. Transfer matrices, locus counts and sharing spectra are descriptive.

Analyses used Python (scikit-learn^44^; lifelines^46^; statsmodels), R (survival; QRISK3 0.6.0; QDiabetes 1.0.2), PLINK 2.0 and LDSC.

## Extended Data figure legends

**Extended Data Fig. 1.**
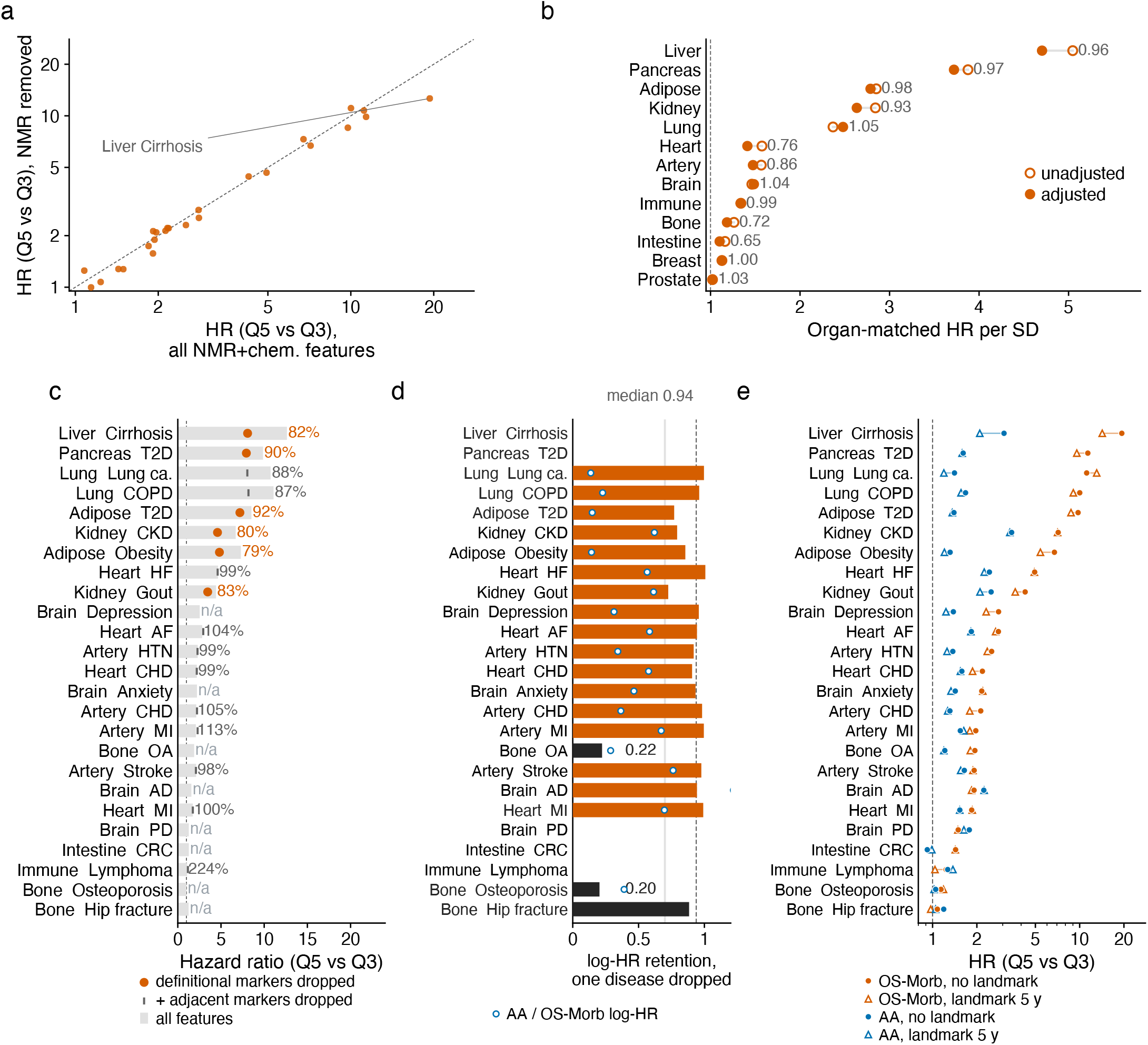
| Robustness of OS-Morb. Multi-omics cohort, n = 22,689; **b**, NMR + chemistry cohort, n = 390,087. **a**, Q5 versus Q3 hazard ratios of the 25 pairs before and after removing all NMR features; the pair with the largest change is labelled. **b**, Organ-matched hazard ratio per SD before and after adding the component shared by the OS-Morb clocks as a covariate; numbers, log-hazard-ratio retention. c, Hazard ratios after removing diagnostic markers; grey bars, full panel; circles, defining markers removed; ticks, adjacent markers also removed; numbers, log-hazard-ratio retention; n/a, no diagnostic marker of that disease in the panel. d, Log-hazard-ratio retention when one disease is dropped from the label and the retrained clock predicts that disease (bars); lines, 0.70 reference and overall median; open circles, log hazard ratio of AA on the same pair relative to OS-Morb. Combinations with a full-composite hazard ratio of 1.10 or below are not interpreted. e, Q5 versus Q3 hazard ratios with events in the first 5 years of follow-up excluded and time reset (log axis), against no exclusion.

**Extended Data Fig. 2.**
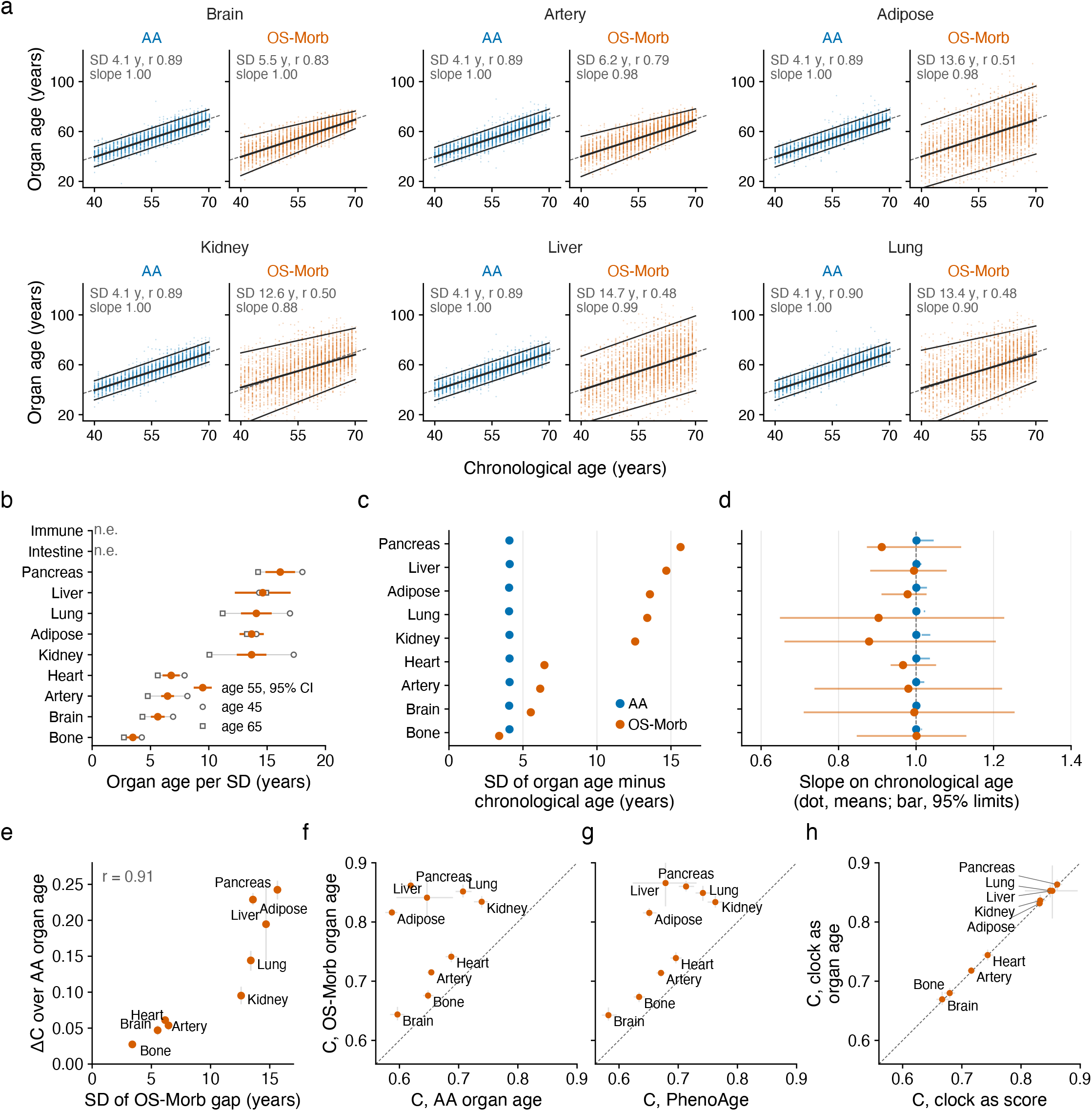
| Organ age in years. a, Organ age against chronological age for the six organs not in Fig. 1c, from the AA clock (chronological age plus its age gap) and from the OS-Morb score converted to years; multi-omics cohort, n = 22,689, 3,000 random people shown; solid lines, mean and 2.5th and 97.5th percentiles in each year of age, each joined by a straight line; dashed line, y = x; SD, standard deviation of organ age minus chronological age; slope, slope of the means on chronological age. b, Organ age per SD of OS-Morb score at ages 45, 55 and 65: the log hazard ratio of the score for first onset of the organ’s own diseases (Cox, adjusted for age and sex, with a score-by-age term) divided by the log hazard ratio of all-cause death per year of age; lines, 95% CI at age 55; n.e., not estimable, the clock did not predict its own diseases. c, Standard deviation of organ age minus chronological age in nine organs, AA and OS-Morb. d, Slope on chronological age of the mean organ age (dots) and of the 2.5th and 97.5th percentile lines (bars); dashed line, 1. e, Standard deviation of the OS-Morb organ-age gap against the gain in Harrell’s C of OS-Morb organ age over AA organ age for first onset of the organ’s own diseases; lines, 95% bootstrap CI. f, Harrell’s C for first onset of each organ’s own diseases with organ age used alone as the predictor: organ age from the OS-Morb score (y) against AA organ age (x). g, As f, against PhenoAge (x); people with PhenoAge, n = 20,298. h, Harrell’s C of one Cox model adjusted for age and sex, with the clock entered as the standardized score (x) or as the organ age gap in years (y). f–h, dashed line, equal C; lines, 95% bootstrap CI; immune system and intestine not converted to years.

**Extended Data Fig. 3.**
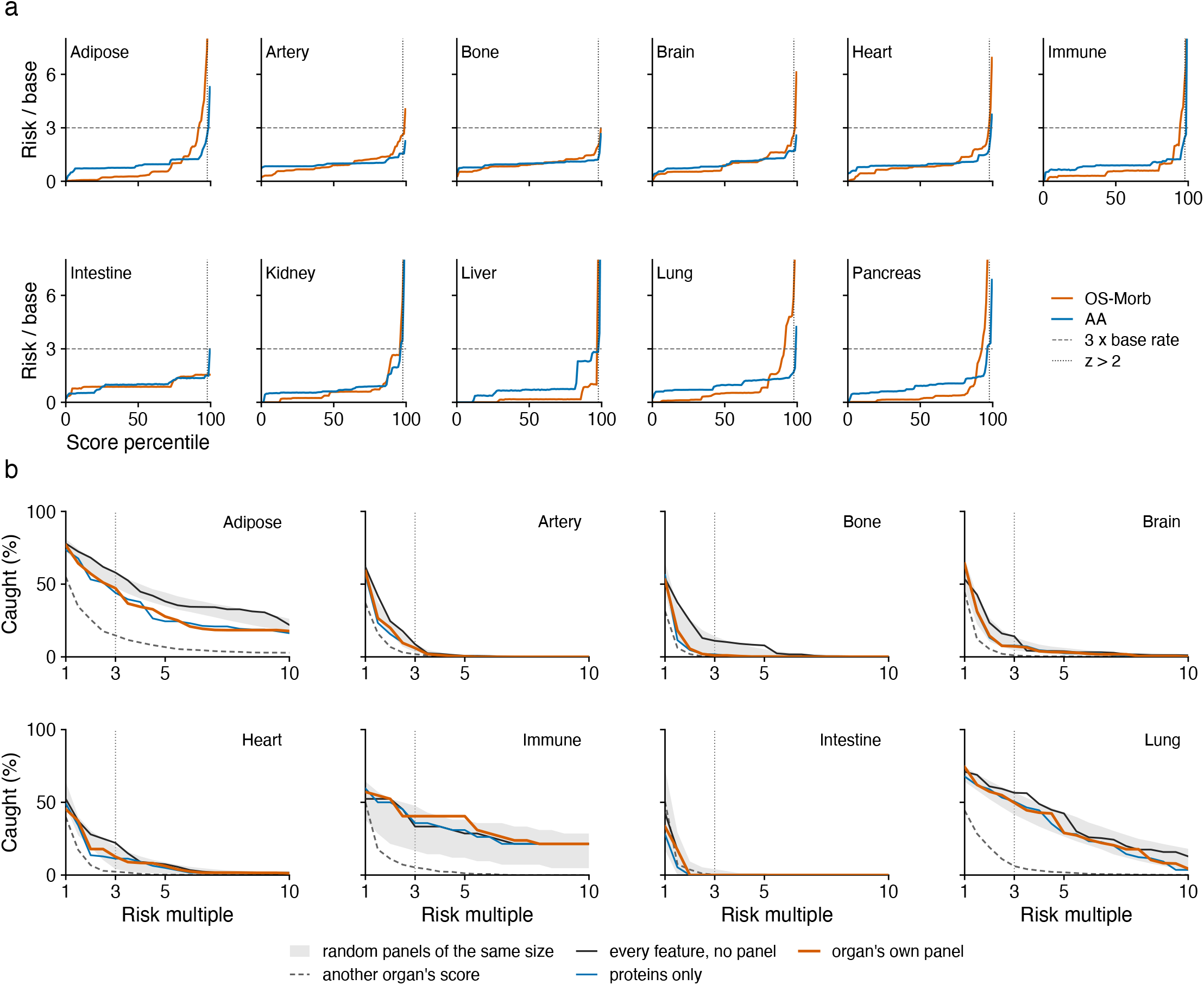
| Risk flags in all organs. Multi-omics cohort, n = 22,689. **a**, Organ-by-organ version of Fig. 4c,d; the two lines are the incidence and age labels. **b**, The eight organs not shown in Fig. 4e; lines and shading as in Fig. 4e.

**Extended Data Fig. 4.**
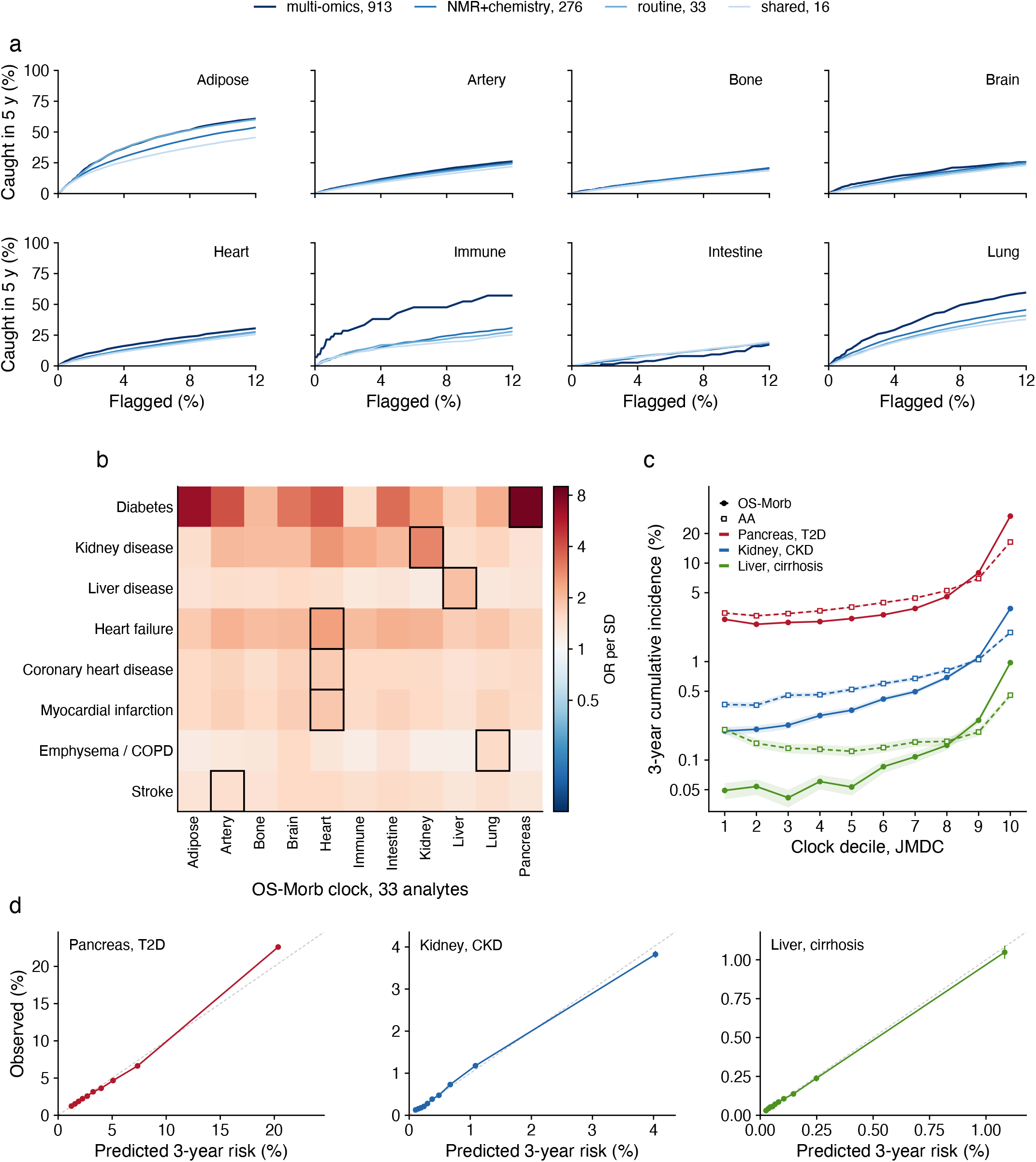
| Capture curves on four substrates, and readouts of routine-test organ clocks in NHANES and JMDC. **a**, Proportion flagged and proportion captured on four substrates for the eight organs not shown in Fig. 5d; multi-omics n = 22,689, other three n = 390,087. **b**, NHANES, n = 48,916. Odds ratios per SD of 11 frozen, transferred OS-Morb organ clocks (33-test panel) for 8 self-reported diagnosed conditions at baseline; logistic regression adjusted for age and sex; boxes, the clock of the organ corresponding to each condition. **c**, JMDC: 3-year cumulative incidence by decile of the pancreas, kidney and liver clock scores, OS-Morb and AA. **d**, JMDC: predicted and observed 3-year incidence by decile of predicted risk for the pancreas, kidney and liver pairs; model of age, sex and the clock score (restricted cubic spline), five-fold out-of-fold predictions; bars, 95% CI of the observed value.

**Extended Data Fig. 5.**
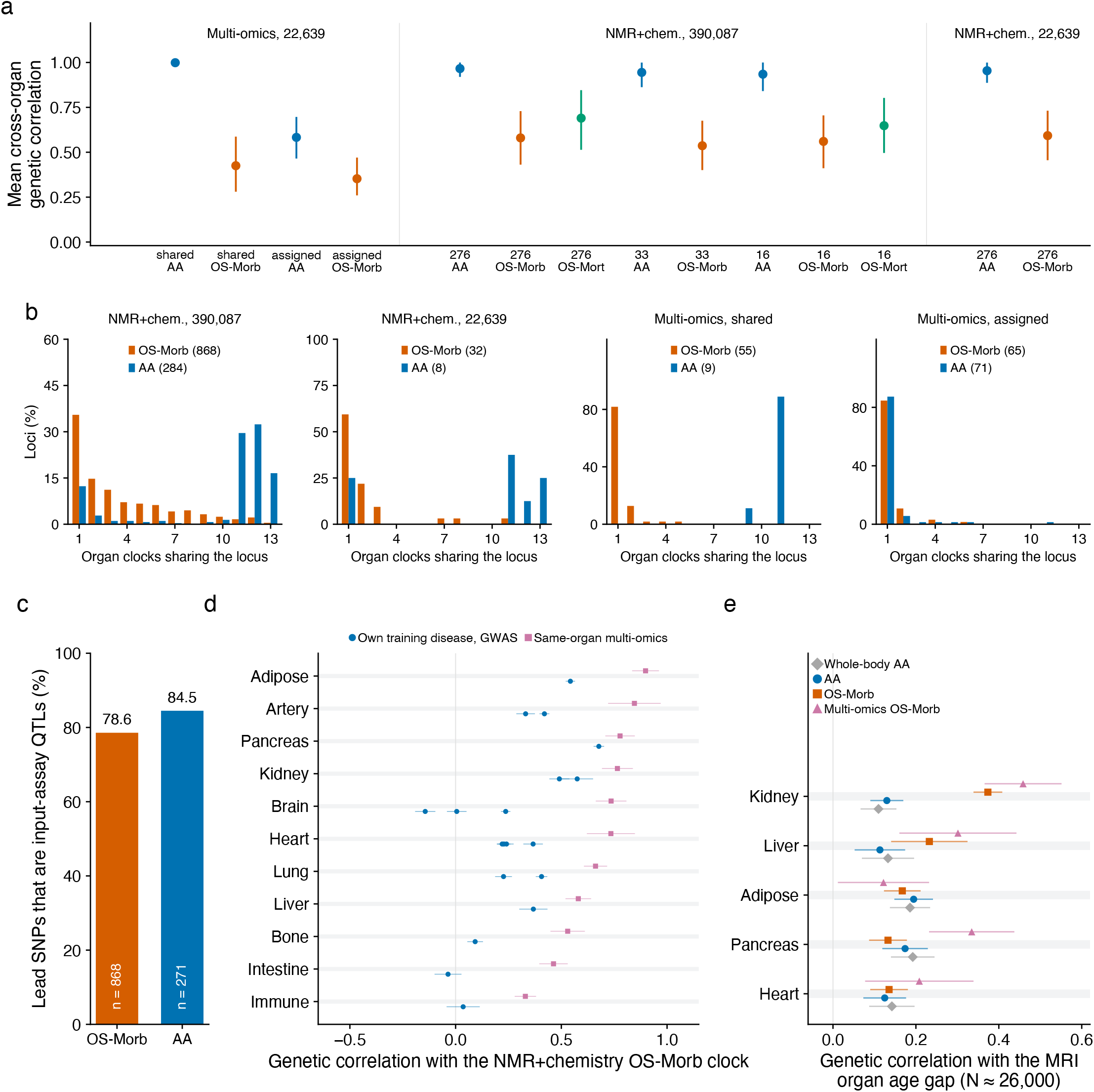
| The training label and the substrate each lower cross-organ genetic correlation. **a**, Mean cross-organ genetic correlation with 95% CI for each GWAS combination (LDSC; bootstrap over organs), grouped by cohort. **b**, Distribution of genome-wide significant loci by the number of clocks hitting them (1 to 13), AA against OS-Morb, loci counts in brackets, y axes differ between panels. The four panels are the full NMR + chemistry cohort, the same features in 22,639 people, shared features in the multi-omics cohort, and assigned protein panels in the multi-omics cohort. **c**, Proportion of lead SNPs that are themselves genome-wide significant QTLs of an input test, full NMR + chemistry cohort, OS-Morb and AA. **d**, Genetic correlation of each NMR + chemistry OS-Morb clock with the GWAS of its own training diseases and with the same-organ multi-omics OS-Morb clock; lines, standard errors. **e**, Genetic correlation of four kinds of clock with MRI organ age gaps^74^, five comparable organs; lines, standard errors.

**Extended Data Fig. 6.**
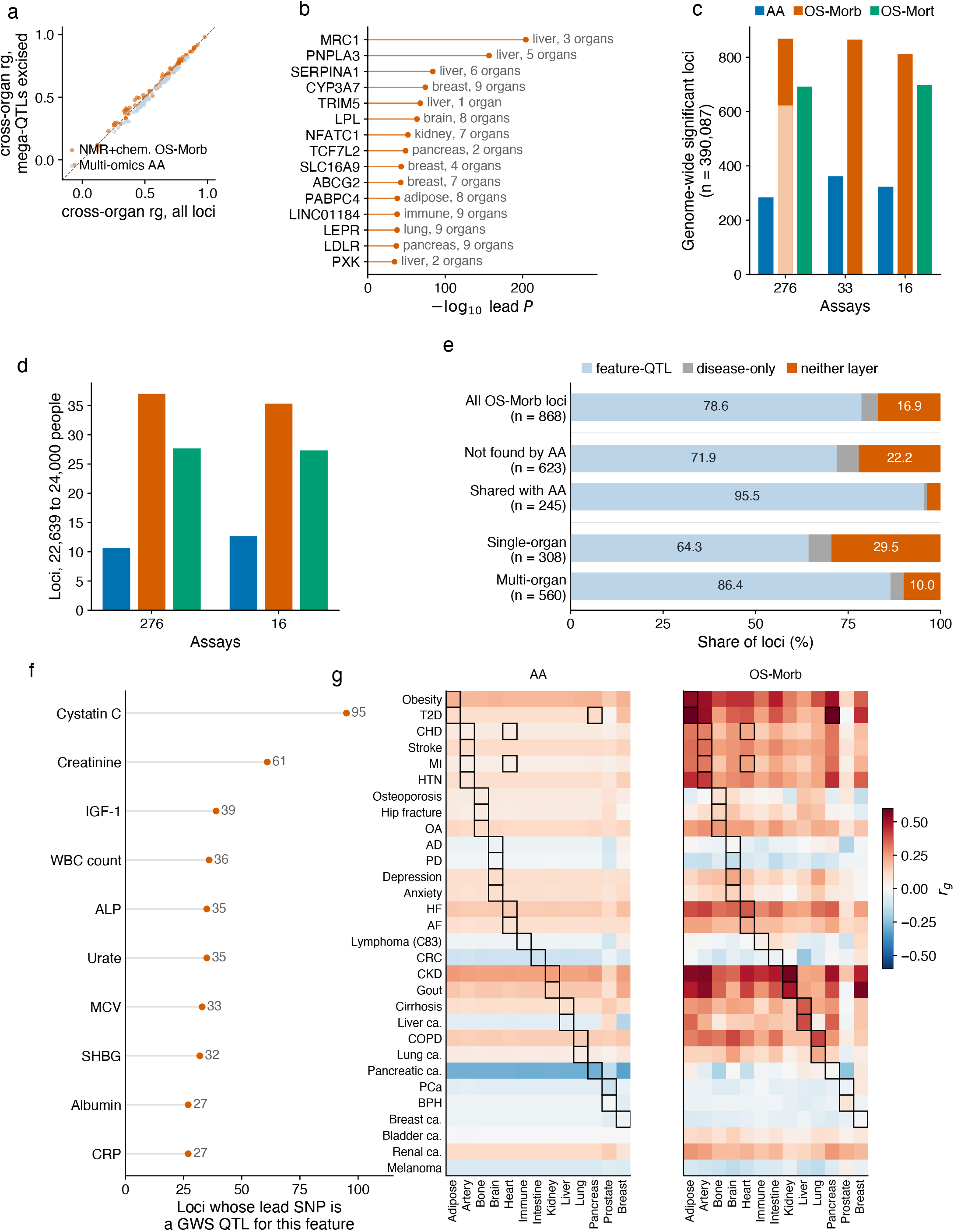
| Supporting genetic panels. **a**, Pairwise cross-organ genetic correlation before and after removing six super-loci regions. **b**, The top 15 OS-Morb-specific loci by lead P value, with nearest gene, leading organ and number of clocks hit. **c**, Number of deduplicated genome-wide significant loci for each substrate × label combination in the NMR + chemistry cohort, n = 390,087, all clocks sharing all features; light segment of the 276-test OS-Morb bar, loci detected by neither AA clock. **d**, The same combinations in 22,639 to 24,000 people. **e**, Three-layer composition of each lead SNP (feature-QTL, disease-only, neither; definitions in Methods); five rows for all loci and two orthogonal splits. **f**, Top ten features by count within the feature-QTL layer. **g**, Genetic correlation of 13 NMR + chemistry clocks with 30 external disease GWAS; left, AA; right, OS-Morb; boxes, each organ’s own diseases.

## Acknowledgements

This research was conducted using UK Biobank data under Application Number 66813. We thank Professor Francisco Lai (Department of Pharmacology and Pharmacy, LKS Faculty of Medicine, The University of Hong Kong) and Dr Deliang Yang (Department of Medicine, School of Clinical Medicine, LKS Faculty of Medicine, The University of Hong Kong) for support with JMDC data access and management.

Large language models were used under full author supervision: Claude (Anthropic) for language editing and analysis-code development, and Gemini (Google) for language editing. All study design, analyses, results and conclusions were conceived, verified and approved by the authors, who take complete responsibility for the content.

## Data Availability

UK Biobank data are available to approved researchers through the UK Biobank Access Management System. NHANES data and linked mortality files are publicly available from the National Center for Health Statistics. JMDC data are available from JMDC Inc. under licence. FinnGen release 10 summary statistics are publicly available. Code to train the organ clocks and apply the frozen coefficients is in a private GitHub repository (https://github.com/shichang855-creator/organ-aging-clocks, folder ms2); access will be given on request during peer review, and the repository will be made public on publication.

## Code availability

Code to train the organ clocks and apply the frozen coefficients will be made available on GitHub on publication.

